# Sex-Specific Association Between Genetic Risk of Psychiatric Disorders and Cardiovascular Diseases

**DOI:** 10.1101/2022.10.08.22280805

**Authors:** Jiayue-Clara Jiang, Kritika Singh, Lea K. Davis, Naomi R. Wray, Sonia Shah

**Author notes:** co-first authors (Jiayue-Clara Jiang, Kritika Singh). **Corresponding authors:** Sonia Shah, PhD Address: 306 Carmody Rd, Institute for Molecular Bioscience, The University of Queensland, St Lucia, Australia, 4072, Jiayue-Clara Jiang, PhD Address: 306 Carmody Rd, Institute for Molecular Bioscience, The University of Queensland, St Lucia, Australia, 4072.

## Abstract

**Background:** The lack of research on female-specific risk factors for cardiovascular diseases (CVD) has led to sex-based disparities in cardiovascular health. Though epidemiological studies show increased CVD risks amongst individuals with psychiatric disorders, findings on sex differences in comorbidity have been inconsistent.

**Methods:** This genetic epidemiology study examined the sex-specific association between the genetic risk of three psychiatric disorders [major depression (MD), schizophrenia and bipolar disorder], estimated using polygenic scores (PGS), and risks of three CVDs [atrial fibrillation (AF), coronary artery disease (CAD) and heart failure (HF)] in 345,169 European-ancestry individuals (UK Biobank), with analyses replicated in an independent BioVU cohort (N=49,057). Mediation analysis was conducted to determine whether traditional CVD risk factors could explain any observed sex difference.

**Results:** In UK Biobank, PGS_MD_ was significantly associated with the incident risks of all three CVDs in females after multiple testing correction [hazard ratio (HR)_AF-female_=1.04 (95% CI: 1.02-1.06; p=0.00015); HR_CAD-female_=1.07 (1.04-1.11; p=2.6E-06); HR_HF-female_=1.09 (1.06-1.13; p=9.7E-10)], but not in males. These female-specific associations remained even in the absence of any psychiatric disorder diagnosis or psychiatric medication use. Although mediation analysis demonstrated that the association between PGS_MD_ and CVDs in females was partly mediated by baseline BMI, hypercholesterolemia, hypertension and smoking, these risk factors did not explain the higher risk compared to males. The association between PGS_MD_ and CAD was consistent between females who were pre-menopausal and post-menopausal at baseline (mean baseline age of 44.9 and 60.8 years, respectively), while the association with AF and HF was only observed in the baseline post-menopausal cohort. No significant association with CVD risks was observed for the PGS of schizophrenia or bipolar disorder. The positive association of PGS_MD_ with CAD and HF risk in females was replicated in BioVU, where the corresponding associations in males also reached nominal significance.

**Conclusions:** Genetic predisposition to MD confers a greater risk of CVDs in females versus males, even in the absence of any depression diagnosis. This study warrants further investigation into whether genetic predisposition to depression could be useful for improving cardiovascular risk prediction even in the absence of a depression diagnosis, especially in women.

## Introduction

The idea that mental health is linked to cardiovascular health has long been recognised: William Harvey wrote in 1628 “every affection of the mind that is attended with either pain or pleasure, hope or fear, is the cause of an agitation whose influence extends to the heart” ^1^. Modern epidemiological studies find that collectively, individuals with psychiatric disorders, namely major depression (MD), schizophrenia (SCZ) and bipolar disorder (BD), have a ∼50% higher odds of developing cardiovascular diseases (CVD) ^2^. The increased CVD risk amongst individuals with psychiatric disorders may be attributed to a combination of genetic and non-genetic factors, with the latter including the use of prescribed psychiatric medications ^3^ and the adoption of adverse habits, such as smoking and social isolation ^4^.

There is marked sex difference in both the prevalence and clinical presentation of psychiatric disorders. Depression is twice as prevalent in females than males (lifetime prevalence of 26.1% versus 14.7%) ^5^, with women showing augmented symptom severity ^6^. While the prevalence of SCZ does not differ between sexes, some studies have found that men show a higher incidence (incidence risk ratio = 1.42), earlier age of onset and worse premorbid functioning ^7^. Men also show an earlier onset for BD, although the prevalence is reported to be equal between sexes for bipolar I disorder but higher in women for bipolar II disorder, which is characterised by depressive episodes ^8^. Despite a higher lifetime risk of CVD in men (60.3% versus 55.6% in women at an index age of 45 years) ^9^, CVD is a leading cause of female deaths ^10^, and yet CVD risk in women remains underappreciated and under-researched, leading to under-diagnosis and under-treatment of CVDs in women ^10^.

Few studies have investigated the sex differences in the cardiovascular comorbidity of SCZ and BD, and observational studies have presented inconsistent findings on the sex-specific association between depression and CVD outcomes ^11^. While some observational studies (N≤3,237) have found depression or depressive symptoms to be a risk factor for heart failure (HF) and coronary artery disease (CAD) amongst women but not in men ^12,13^, a study on a Chinese cohort (N=512,712) found an association between depression and CVD-related mortality in men only ^14^. The variability in findings may be due to differences in sample sizes, unmeasured confounders, follow-up times, sex balance in the cohort, and the criteria for defining depression phenotypes and CVD outcomes. At the same time, observational studies cannot establish causal associations and cannot determine if this risk is a direct result of medication, which are known to have an adverse cardiometabolic effect ^3^, or other environmental factors in consequence of a diagnosis of depression. Furthermore, although an association between depressive symptoms and increased CAD risk in women aged ≤ 55 years (but not in women aged > 55 years) was previously observed ^13^, it remains unknown how the menopause transition (perimenopause), a period where adverse changes in body composition, lipids, and measures of vascular health occur ^15^, affects the cardiovascular comorbidity amongst women with a higher risk of depression.

Large genome-wide association studies (GWAS) have been very successful at identifying genetic loci associated with disease risk. Novel statistical methods applied to GWAS data are facilitating our understanding of shared biology between diseases, as well as in making causal inferences in disease comorbidity, overcoming some of the caveats of observational studies. Previous genetic analyses, for example, have used Mendelian randomisation (MR) to provide evidence to support a causal effect of depression on CAD and HF ^16^. However, few studies have investigated sex differences using such genetic approaches, with an example being a small-scale study (N=18,385, 50.9% female) that showed a positive association between MD and CAD in females but not males ^17^. To what extent genetic factors contribute to comorbidity between psychiatric disorders and CVDs in different sexes is an outstanding question, and one that will better inform CVD prevention strategies. In this study, we used polygenic scores (PGS) derived from GWAS summary statistics (which estimate an individual’s genetic liability to a disease) to examine the sex-specific association between a higher genetic risk of three psychiatric disorders [BD, MD and SCZ] and the risk of three CVDs [atrial fibrillation (AF), CAD and HF] in the UK Biobank cohort of over 345,000 individuals. The findings were independently replicated in a BioVU cohort of over 49,000 individuals.

## Methods

### UK Biobank cohort summary

The UK Biobank cohort consists of over 500,000 participants (aged 40-69 years at baseline) from the UK, who were recruited between 2006 and 2010 ^18^. Diverse phenotypic and genomic data were collected from participants, such as clinical diagnosis, physical and biochemical measurements, and answers from questionnaires. Consent was collected from all participants. The current study was conducted using the UK Biobank Resource under Application Number 12505. This research is covered by The University of Queensland Human Research Ethics Committee approval (HREC number 2020/HE002938).

Using information provided by the UK Biobank genotype data release, quality control was performed to remove individuals who were outliers for heterozygosity and genotype missing rates, who were excluded from kinship inference, who showed evidence of putative sex chromosome aneuploidy, and who showed mismatching self-reported and genetically inferred sex (definitions of exclusion criteria are described in Bycroft et al. ^18^). Individuals who have withdrawn from the study or have missing phenotypes [baseline body mass index (BMI) or smoking status] were excluded from the analysis. After quality control, a total of 345,169 unrelated (genetic relatedness < 0.05) individuals of European ancestry (inferred from the genetic data) were subjected to analysis in this study (ancestry calling is described in Yengo et al. ^19^).

### BioVU cohort summary

Vanderbilt University Medical Center (VUMC) is a tertiary care centre that provides inpatient and outpatient care in middle Tennessee, US. The VUMC electronic health record (EHR) system, established in 1990, includes data on billing codes from the International Classification of Diseases 9^th^ and 10^th^ editions (ICD-9 and ICD-10), Current Procedural Terminology (CPT) codes, laboratory values, reports, and clinical documentation. In 2007, VUMC launched a biobank, BioVU, which links a patient’s DNA sample to their EHR. The BioVU consent form is provided to patients in the outpatient clinic environments at VUMC. The VUMC Institutional Review Board oversees BioVU and approved this project (IRB#172020). At VUMC, all medical record data were extracted from the EHRs of 72,634 individuals of European ancestry, determined by genetic ancestry analysis. To reduce missingness of clinical data and enrich the sample for patients who receive their primary care at VUMC, our analysis included only individuals who met the “medical home” definition, requiring the presence of any five codes on different days over a period of at least three years. We further excluded individuals who did not have a BMI measurement or age recorded in the EHRs, leaving a total of 49,057 individuals for analysis.

### UK Biobank phenotype ascertainment

UK Biobank phenotypic data collected between 2006 and March 2023 was used for phenotype extraction. Detailed inclusion and exclusion criteria of all disease phenotypes in the UK Biobank are provided in Supplementary Methods, Supplementary Table 1 and Supplementary Table 2.

For each CVD, prevalent cases consisted of individuals who had disease-associated phenotypes recorded at baseline or after recruitment into UK Biobank, defined using self-reported illnesses (data-field 20002), self-reported operation codes (data-field 20004), operative procedures (OPCS4) (data-field 41272), ICD-9 diagnoses (data-field 41271), ICD-10 diagnoses (data-field 41270) and primary (data-field 40001) or contributory (data-field 40002) death causes (not all criteria were used for all phenotype definitions). Incident cases were defined as individuals who did not self-report any disease-associated illness or operation code at initial assessment [determined using data-field 53 (“Date of attending assessment centre”), data-field 20008 (“Interpolated Year when non-cancer illness first diagnosed”) and data-field 20010 (“Interpolated Year when operation took place”)], and did not have any disease-associated OPCS4 procedure or diagnosis (ICD-9 or ICD-10) prior to or at the time of initial assessment [determined using data-field 41282 (“Date of first operative procedure - OPCS4”), data-field 41281 (“Date of first in-patient diagnosis - ICD-9”) and data-field 41280 (“Date of first in-patient diagnosis - ICD-10”)]. For prevalent cases who answered “Date uncertain or unknown” or “Preferred not to answer” to data-fields 20008 and 20010, the corresponding self-reported illness diagnosis or operation date was set to be the date on which they attended the assessment centre when they self-reported the illness or operation (data-field 53). The process of defining prevalent and incident cases is presented in Figure 1.

**Figure 1.**
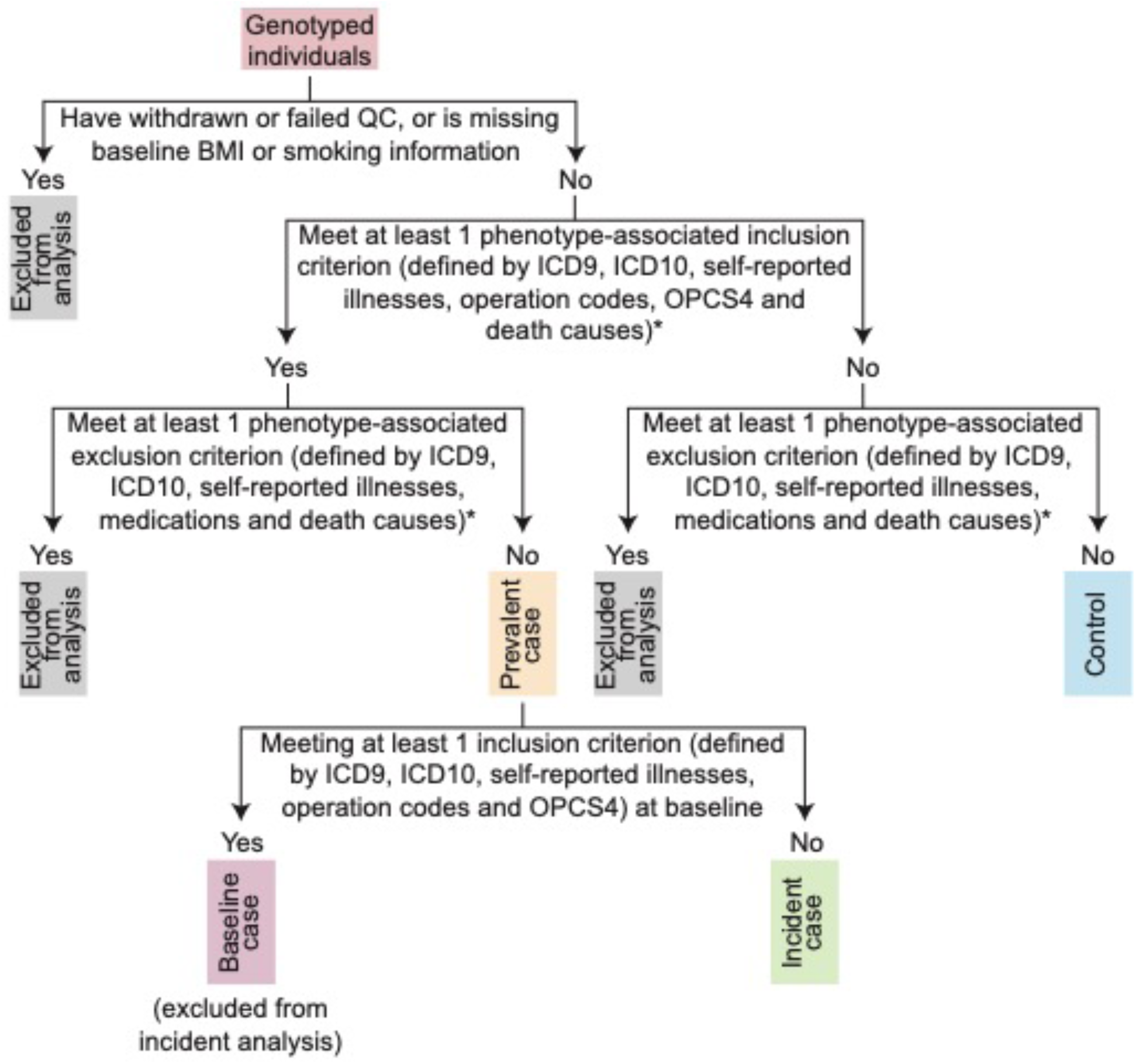
Flow chart for defining prevalent and incident CVD cases in the UK Biobank cohort. *The full inclusion and exclusion criteria for each disease are presented in Supplementary Table 1.

For the mediation analysis, the baseline status of the disease risk factors of CVDs [hypercholesterolemia, hypertension, and type II diabetes (T2D)] was defined using criteria in Supplementary Table 1. Incident cases of these risk factors, defined as described above, were removed from the mediation analysis.

Smoking status at baseline (whether a person has smoked ever, defined as current or previous smokers) was collated from UK Biobank data-field 20116 (“Smoking status”). The baseline BMI of individuals was retrieved from data-field 21001 [“Body mass index (BMI)”]. The ages at baseline (in years) of individuals were calculated using “Date of attending assessment centre” (data-field 53) and “Year of birth” (data-field 34). Summary of characteristics for the whole cohort and the cohort with no psychiatric diagnosis and medications are shown in Table 1 and Supplementary Table 3 respectively.

**Table 1.**
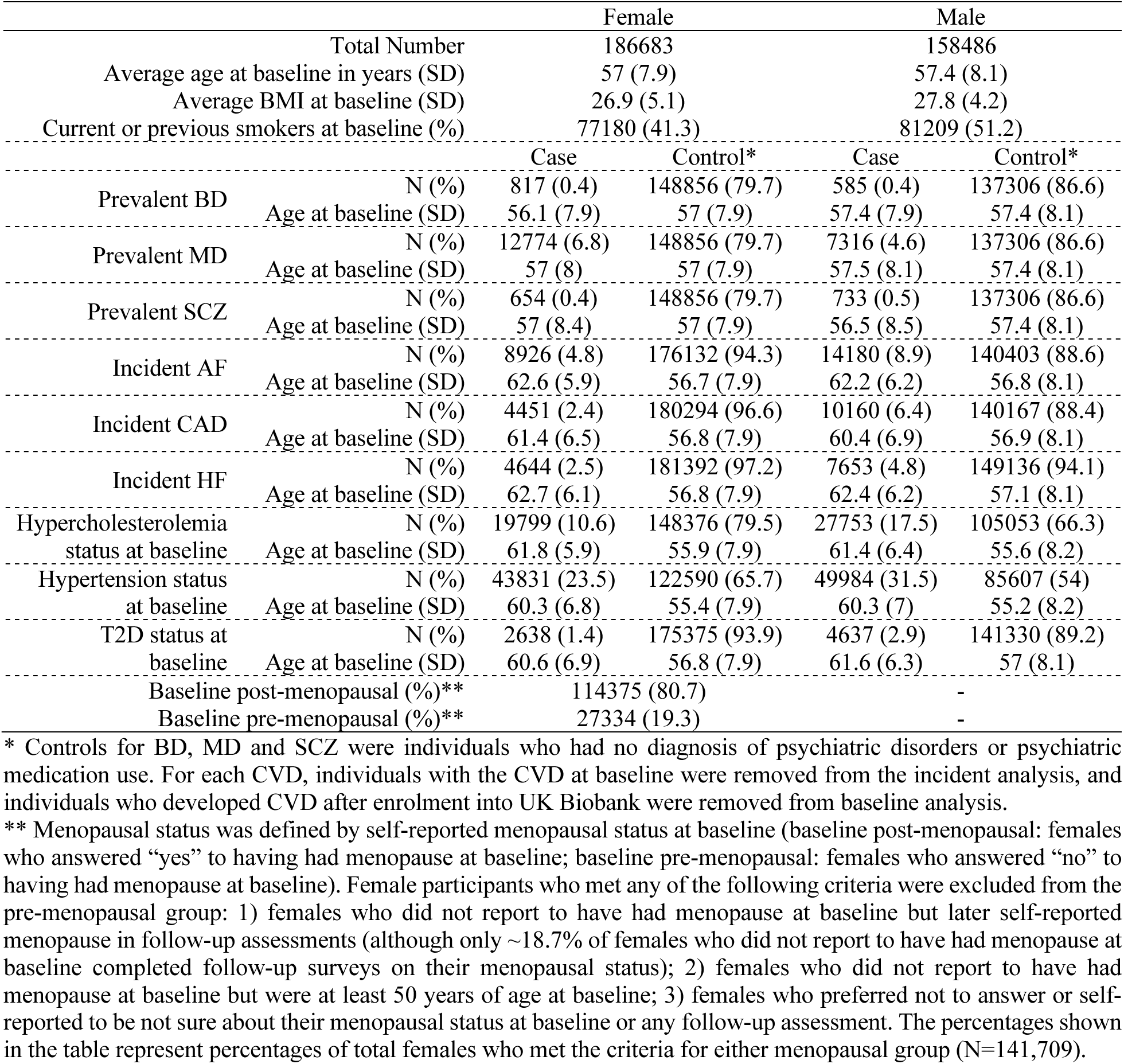
Summary of characteristics of the UK Biobank cohort.

|  |  | Female |  | Male |  |
| --- | --- | --- | --- | --- | --- |
| Total Number |  | 186683 |  | 158486 |  |
| Average age at baseline in years (SD) |  | 57 (7.9) |  | 57.4 (8.1) |  |
| Average BMI at baseline (SD) |  | 26.9 (5.1) |  | 27.8 (4.2) |  |
| Current or previous smokers at baseline (%) |  | 77180 (41.3) |  | 81209 (51.2) |  |
|  |  | Case | Control* | Case | Control* |
| Prevalent BD | N (%) | 817 (0.4) | 148856 (79.7) | 585 (0.4) | 137306 (86.6) |
|  | Age at baseline (SD) | 56.1 (7.9) | 57 (7.9) | 57.4 (7.9) | 57.4 (8.1) |
| Prevalent MD | N (%) | 12774 (6.8) | 148856 (79.7) | 7316 (4.6) | 137306 (86.6) |
|  | Age at baseline (SD) | 57 (8) | 57 (7.9) | 57.5 (8.1) | 57.4 (8.1) |
| Prevalent SCZ | N (%) | 654 (0.4) | 148856 (79.7) | 733 (0.5) | 137306 (86.6) |
|  | Age at baseline (SD) | 57 (8.4) | 57 (7.9) | 56.5 (8.5) | 57.4 (8.1) |
| Incident AF | N (%) | 8926 (4.8) | 176132 (94.3) | 14180 (8.9) | 140403 (88.6) |
|  | Age at baseline (SD) | 62.6 (5.9) | 56.7 (7.9) | 62.2 (6.2) | 56.8 (8.1) |
| Incident CAD | N (%) | 4451 (2.4) | 180294 (96.6) | 10160 (6.4) | 140167 (88.4) |
|  | Age at baseline (SD) | 61.4 (6.5) | 56.8 (7.9) | 60.4 (6.9) | 56.9 (8.1) |
| Incident HF | N (%) | 4644 (2.5) | 181392 (97.2) | 7653 (4.8) | 149136 (94.1) |
|  | Age at baseline (SD) | 62.7 (6.1) | 56.8 (7.9) | 62.4 (6.2) | 57.1 (8.1) |
| Hypercholesterolemia status at baseline | N (%) | 19799 (10.6) | 148376 (79.5) | 27753 (17.5) | 105053 (66.3) |
|  | Age at baseline (SD) | 61.8 (5.9) | 55.9 (7.9) | 61.4 (6.4) | 55.6 (8.2) |
| Hypertension status at baseline | N (%) | 43831 (23.5) | 122590 (65.7) | 49984 (31.5) | 85607 (54) |
|  | Age at baseline (SD) | 60.3 (6.8) | 55.4 (7.9) | 60.3 (7) | 55.2 (8.2) |
| T2D status at baseline | N (%) | 2638 (1.4) | 175375 (93.9) | 4637 (2.9) | 141330 (89.2) |
|  | Age at baseline (SD) | 60.6 (6.9) | 56.8 (7.9) | 61.6 (6.3) | 57 (8.1) |
| Baseline post-menopausal (%)** |  | 114375 (80.7) |  | - |  |
| Baseline pre-menopausal (%)** |  | 27334 (19.3) |  | - |  |

\* Controls for BD, MD and SCZ were individuals who had no diagnosis of psychiatric disorders or psychiatric medication use. For each CVD, individuals with the CVD at baseline were removed from the incident analysis, and individuals who developed CVD after enrolment into UK Biobank were removed from baseline analysis.
\*\* Menopausal status was defined by self-reported menopausal status at baseline (baseline post-menopausal: females who answered “yes” to having had menopause at baseline; baseline pre-menopausal: females who answered “no” to having had menopause at baseline). Female participants who met any of the following criteria were excluded from the pre-menopausal group: 1) females who did not report to have had menopause at baseline but later self-reported menopause in follow-up assessments (although only ~18.7% of females who did not report to have had menopause at baseline completed follow-up surveys on their menopausal status); 2) females who did not report to have had menopause at baseline but were at least 50 years of age at baseline; 3) females who preferred not to answer or self-reported to be not sure about their menopausal status at baseline or any follow-up assessment. The percentages shown in the table represent percentages of total females who met the criteria for either menopausal group (N=141,709).

### BioVU phenotype ascertainment

For the BioVU cohort, we defined phenotypes based on Phecodes, which are higher-order combinations of at least two related ICD codes, occurring on two different days, using the R PheWAS package ^20^ (detailed Phecodes are shown in Supplementary Table 4). Smoking status was ascertained by “tobacco use disorder” (Phecode 318). The median BMI and median age of individuals were selected from assessments taken at chronological visits, available in the medical records at VUMC. Summary of characteristics is presented in Supplementary Table 5.

### GWAS summary statistics

Association summary statistics derived from European-ancestry or predominantly European-ancestry cohorts were retrieved for three CVDs (AF, CAD and HF) and three psychiatric disorders (BD, MD and SCZ), and were used for generating PGS in the UK Biobank and BioVU cohorts (datasets summarised Supplementary Table 6). To avoid discovery bias, GWAS summary statistics used for single nucleotide polymorphism (SNP) selection and per-allele weight estimation were derived from studies that did not include individuals from the corresponding target cohorts (UK Biobank or BioVU).

### Polygenic score generation

PGS is an estimate of the lifetime genetic liability of an individual to a trait. Briefly, we define a PGS of an individual, *j*, as a weighted sum of SNP allele counts: 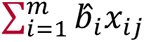, where *m* is the number of SNPs included in the predictor, 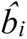 is the per-allele weight (based on the estimated effect of the allele on trait value) for the SNP, *x_ij_* is a count of the number (0, 1, or 2) of trait-associated alleles of SNP *i* in individual *j*.

Several methods are available for optimising the choice of SNPs and the per-allele weights for PGS calculation. For analysis in the UK Biobank cohort, we used SBayesR ^21^ which implements a Bayesian approach to model genetic architecture, is computationally efficient, and does not require a training dataset for identifying trait-associated SNPs and estimating per-allele weights ^21,22^. A linkage disequilibrium (LD) reference dataset was downloaded from the SBayesR website (banded LD matrix) and default settings were used unless otherwise stated (--exclude-mhc; -- chain-length 50000; --burn-in 10000; --no-mcmc-bin) ^21^. For each disease, we applied PLINK1.9 ^23^ to calculate the PGSs (based on SbayesR SNPs and weights) in UK Biobank individuals. For BioVU, PGSs were generated using Polygenic Risk Score–Continuous Shrinkage (PRS-CS) ^24^ (which has been shown to have similar performance to SbayesR ^22^). The LD reference panel was constructed from 503 European samples in the 1000 Genomes Project Phase 3.41.

The PGS distribution was scaled to a mean of zero and standard deviation (SD) of 1; therefore, effect size estimates in subsequent regression analyses were interpreted as changes in outcome per 1-SD increase in PGS. We generated PGS for 345,169 unrelated, European-ancestry individuals [186,683 (54.1%) females and 158,486 (45.9%) males] from the UK Biobank, and 49,057 individuals [28,094 (57.3%) females and 20,963 (42.7%) males] from BioVU. To validate the PGS, logistic regression was used to examine the association with disease prevalent cases in the prediction cohort (UK Biobank or BioVU) (Supplementary Table 7). PGS performance was evaluated using the Nagelkerke pseudo R^2^ [R package “rcompanion” ^25^ (version 2.4.13)]. The Nagelkerke pseudo R^2^ compares the reduced model [for UK Biobank: containing only sex, BMI, smoking status, genotyping array, age at baseline and 20 genetic principal components (PC) as predictors; for BioVU: containing only sex, BMI, smoking status, age and 10 genetic PCs as predictors] and the full model (with PGS added as a predictor), and is a relative measure of how well the full model explains the data compared to the reduced model. Overall, the addition of PGSs improved the prediction performance in our study, as indicated by positive Nagelkerke R^2^ values (Supplementary Table 7). Our models demonstrate comparable performance to the corresponding PGSs generated previously using different methods ^24,26–28^ (Supplementary Table 7).

### Cox proportional hazards regression analysis

In UK Biobank, for each psychiatric-CVD disorder pair, we performed age-as-time-scale Cox proportional hazards regression to investigate the sex-specific association between psychiatric disorder PGS and incident CVD. For each CVD incident case, we defined their time-to-event as the number of months since birth [estimated using “Year of birth” (data-field 34) and “Month of birth” (data-field 52)], to the age they were diagnosed with CVD [estimated using “Date of first in-patient diagnosis” (data-field 41280)], first self-reported CVD illness [estimated using “Interpolated Year when non-cancer illness first diagnosed” (data-field 20008)], self-reported operation codes [estimated using “Interpolated Year when operation took place” (data-field 20010)], had the CVD-associated OPCS4 procedures [estimated using “Date of first operative procedure - OPCS4” (data-field 41282)] or died of CVDs [estimated using “Date of death” (data-field 40000)]. If an individual met multiple disease-associated criteria (such as having had both a disease-associated ICD-10 diagnosis and OPCS4 procedure), their time-to-event was defined as the earliest time at which they met any disease-associated criteria.

We censored CVD controls at the age of death, age of losing contact [estimated using “Date lost to follow-up” (data-field 191)], or age at censoring (March 2023). The Cox models included BMI, smoking status, genotyping array and 20 genetic PCs as covariates, and were analysed using the “Survival” package (version 3.2.11) in R ^29^. A p<0.0028 was chosen to indicate statistical significance (multiple testing correction for 18 tests – association of three psychiatric PGS with three CVDs in two sexes). The regression coefficients (beta) for the PGS were compared between sexes using a Wald test (statistical significance declared at two-sided p<0.05). Given previously reported genetic correlations between CVD and depression ^30^, to assess if the MD PGS were associated with CVD risks independently of the CVD PGS, we performed a sensitivity analysis where the PGS for the relevant CVD was fitted as an additional covariate. Due to the bidirectional relationships amongst the different CVDs, we performed a further sensitivity analysis where the PGSs for all CVDs were fitted as additional covariates. For all subsequent subgroup analyses, a nominal significance threshold (p<0.05) was applied due to the reduction in sample sizes.

### Cox proportional hazards analysis in females stratified by baseline menopausal status

We sought to determine if the association between genetic risk of MD and risk of incident CVD differed between two groups of women at different stages of menopause at baseline. Menopause status at baseline was determined using UK Biobank data-field 2724 (“Had menopause?”). We first explored the MD-CVD association amongst females who were post-menopausal at baseline (who answered “yes” to the survey question “Had menopause?” at the initial assessment visit). The mean age of natural menopause is 50 years (interquartile range = 48.0–53.0 years) ^15^, we thus also conducted analysis in a cohort consisting of females who at baseline were less than 50 years of age and pre-menopausal (answered “no” to “Had menopause?” at the initial assessment), and either answered “no” or did not answer the “Had menopause” question in subsequent assessments (about 18.7% of females who answered “no” to “Had menopause?” at the initial assessment completed at least one follow-up assessment on menopausal status). Females who at baseline did not answer, answered “preferred not to answer”, or answered “not sure” to the menopause question were excluded from both menopausal groups, while those who in any follow-up assessments reported to have had menopause, answered “preferred not to answer”, or answered “not sure” were also excluded from the baseline pre-menopausal group.

A total of 141,709 females in the study met the criteria for either menopausal group, including 114,375 (80.7%) females who self-reported to have had menopause at baseline, and 27,334 (19.3%) females in the baseline pre-menopausal group. We performed Cox proportional hazards regression analysis (as described above) between PGS_MD_ and incident CVDs in the two menopausal groups. BMI, smoking status, genotyping array, and 20 genetic PCs were included in the model as covariates. For each CVD, we applied a Wald test to compare the Cox regression coefficient (beta) for PGS_MD_ in each menopausal group against the previously estimated coefficient in males.

### Mediation analysis with CVD-associated risk factors

To explore whether the association between the genetic risk of MD and incident CVDs can be explained by a CVD-associated risk factor, sex-stratified mediation analysis was performed using the “mediation” R package (version 4.5.0) ^31^. Five risk factors (baseline BMI, hypercholesterolemia status at baseline, hypertension status at baseline, smoking status at baseline and T2D status at baseline) were modelled as mediators between PGS_MD_ (exposure) and incident CVD (outcome). The mediation models included baseline BMI (except in the analysis of baseline BMI as the mediator), baseline smoking status (except in the analysis of smoking status as the mediator), age at baseline, genotyping array and 20 genetic PCs as covariates. Each mediation analysis was performed with 10,000 bootstrap simulations to estimate the variance and significance of the mediation, and statistical significance was defined by p<0.0017 (multiple testing correction for three CVDs, five risk factors and two sexes).

### Association between the genetic risk of MD and CVD in individuals with no diagnosis of psychiatric disorders or psychiatric medication use

To dissociate the effects of behavioural changes or medication use as a consequence of depression diagnosis, sex-stratified Cox proportional hazards regression analysis was performed between PGS_MD_ and incident CVD risks amongst UK Biobank participants who did not meet the criteria for the three psychiatric disorders, had no ICD diagnosis of personality or neurotic disorders, had no self-reported history of mood disorders, and were not on any antidepressants or antipsychotic medications (detailed phenotype information in Supplementary Table 2). A total of 286,162 individuals from the UK Biobank cohort met the above criteria and were included for analysis.

### Logistic regression in BioVU

Findings from UK Biobank were tested for association in the independent BioVU cohort at VUMC. Based on methods used in previous BioVU studies ^17^, logistic regression was used to estimate the sex-stratified association of psychiatric disorder PGS with prevalent CVDs, including median age, median BMI, smoking status and 10 genetic PCs as covariates. In a sensitivity analysis, we additionally adjusted for the presence of a Phecode for any of the psychiatric conditions (BD, MD, SCZ, personality disorders, mood disorders, or anxiety, dissociative and somatoform disorders) (Supplementary Table 4), along with the presence of antidepressant use, defined as whether a person has ever used an antidepressant. Statistical significance was declared at p<0.05.

## Results

### Increased genetic risk of MD is associated with a greater increase of AF, CAD and HF risk in females

Sex-stratified Cox proportional hazards regression analysis showed that after multiple testing correction (p<0.0028), a 1-SD increase in PGS_MD_ was significantly associated with increased risks of all three CVDs in females [hazard ratio (HR)_AF-female_ = 1.04 (95% CI: 1.02 - 1.06; p=0.00015); HR_CAD-female_ = 1.07 (95% CI: 1.04 - 1.11; p=2.6E-06); HR_HF-female_ = 1.09 (95% CI: 1.06 - 1.13; p=9.7E-10)], but not in males, and the association with incident CAD and HF was significantly higher in females compared to males (two-sided Wald test p=0.016 and 0.00032 respectively) (Figure 2). These associations remained even after correcting for the genetic risks of all three CVDs (Supplementary Figure 1), indicating that PGS_MD_ captured additional risk for CVD over and above that captured by the PGSs for all three CVDs. No associations between PGS for SCZ or BD with CVD risks passed multiple testing correction in either sex (Figure 2).

**Figure 2.**
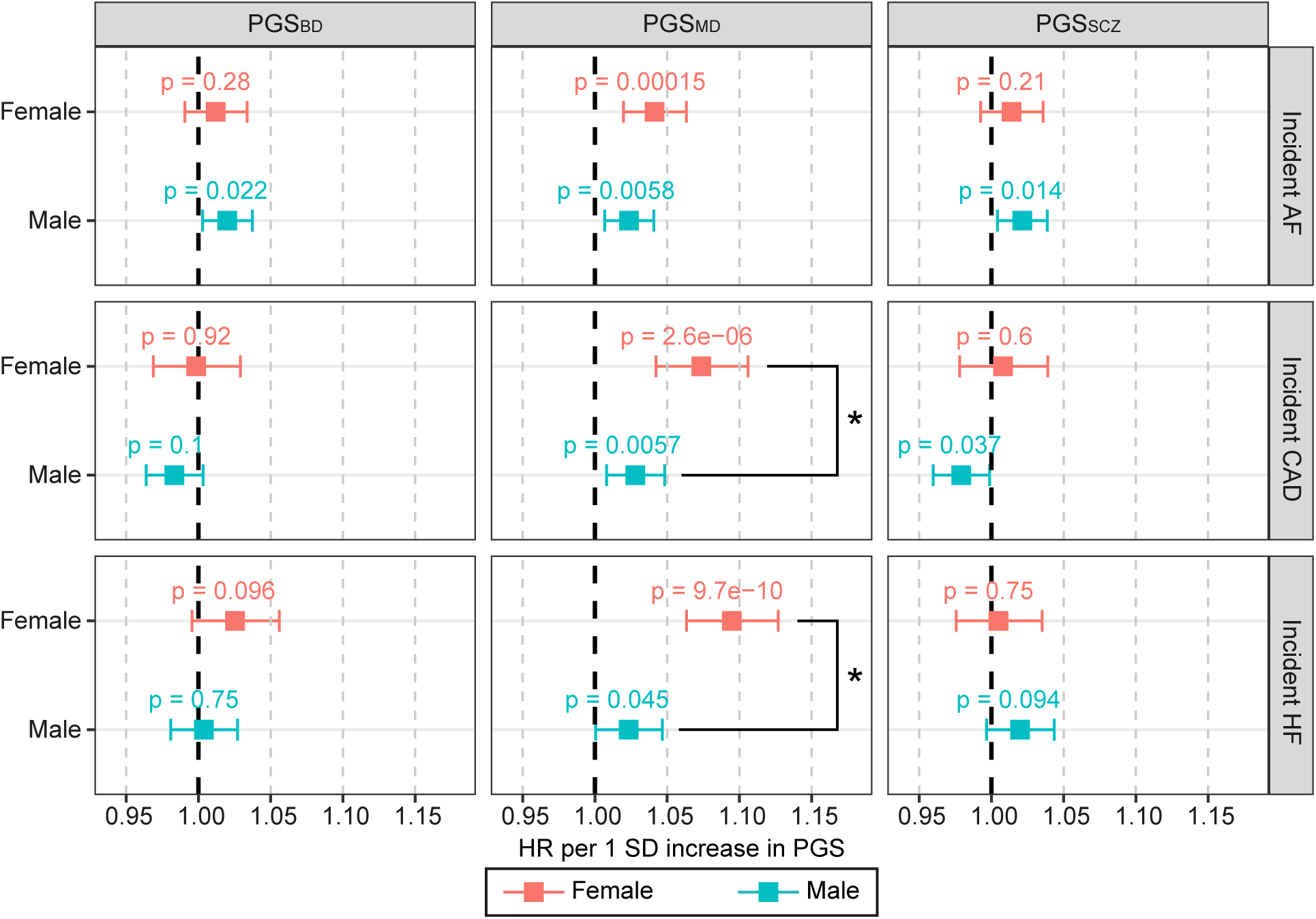
Age-specific change in risk of incident CVDs per SD increase in psychiatric disorder PGSs in the UK Biobank cohort. Associations for the sex-stratified cohorts (red - female; blue - male) were estimated with Cox proportional hazards regression models. X-axis shows the HR per SD increase in PGS, with p-values labelled, and error bars indicate 95% confidence intervals. Asterisk (*) indicates a statistically significant difference in the beta values between females and males (two-sided Wald test p<0.05). Dark grey line indicates HR of 1. AF, atrial fibrillation; BD, bipolar disorder; CAD, coronary artery disease; HF, heart failure; HR, hazard ratio; MD, major depression; PGS, polygenic score; SCZ, schizophrenia; SD, standard deviation.

### Menopausal status influences the association between genetic risk of MD and incident CVDs

We further explored the association between PGS_MD_ and incident CVDs in females stratified by their baseline menopausal status. As expected from previous evidence on the increase in CVD risks after menopause ^15^, the incidence of CVD was higher in the post-menopausal group (6.1%, 2.9% and 3.1% for AF, CAD and HF, respectively), compared to the pre-menopausal group (0.8%, 0.7% and 0.5% for AF, CAD and HF, respectively) (chi square p<2E-16) (Supplementary Table 8). Using a nominal significance (p<0.05) threshold, we found that PGS_MD_ was significantly associated with increased incident CAD regardless of baseline menopausal status [HR_CAD-pre_ = 1.22 (95% CI: 1.06 - 1.42; p=0.0063); HR_CAD-post_ = 1.07 (95% CI: 1.03 - 1.11; p=0.00012)]. The associations with CAD risk in both the baseline pre- and post-menopausal female groups were higher than in males (two-sided Wald test p=0.019 and 0.047, respectively) (Figure 3). A 1-SD increase in PGS_MD_ was also associated with incident AF and HF in the baseline post-menopausal females [HR_AF-post_ = 1.03 (95% CI: 1.01 - 1.06; p=0.0064); HR_HF-post_ = 1.10 (95% CI: 1.06 - 1.13; p=8.2E-08)], and the association with incident HF was significantly higher than the male cohort (two-sided Wald test p=0.00089). Despite risk estimates that were higher than or similar to those observed in the post-menopausal cohort, the associations between PGS_MD_ with incident AF or HF in the baseline pre-menopausal group were not statistically significant [HR_AF-pre_ = 1.07 (95% CI: 0.94 – 1.23; p=0.29; HR_HF-pre_ = 1.10 (95% CI: 0.92 – 1.30; p=0.30)], which may be a reflection of the much smaller sample size for the pre-menopausal cohort (Figure 3).

**Figure 3.**
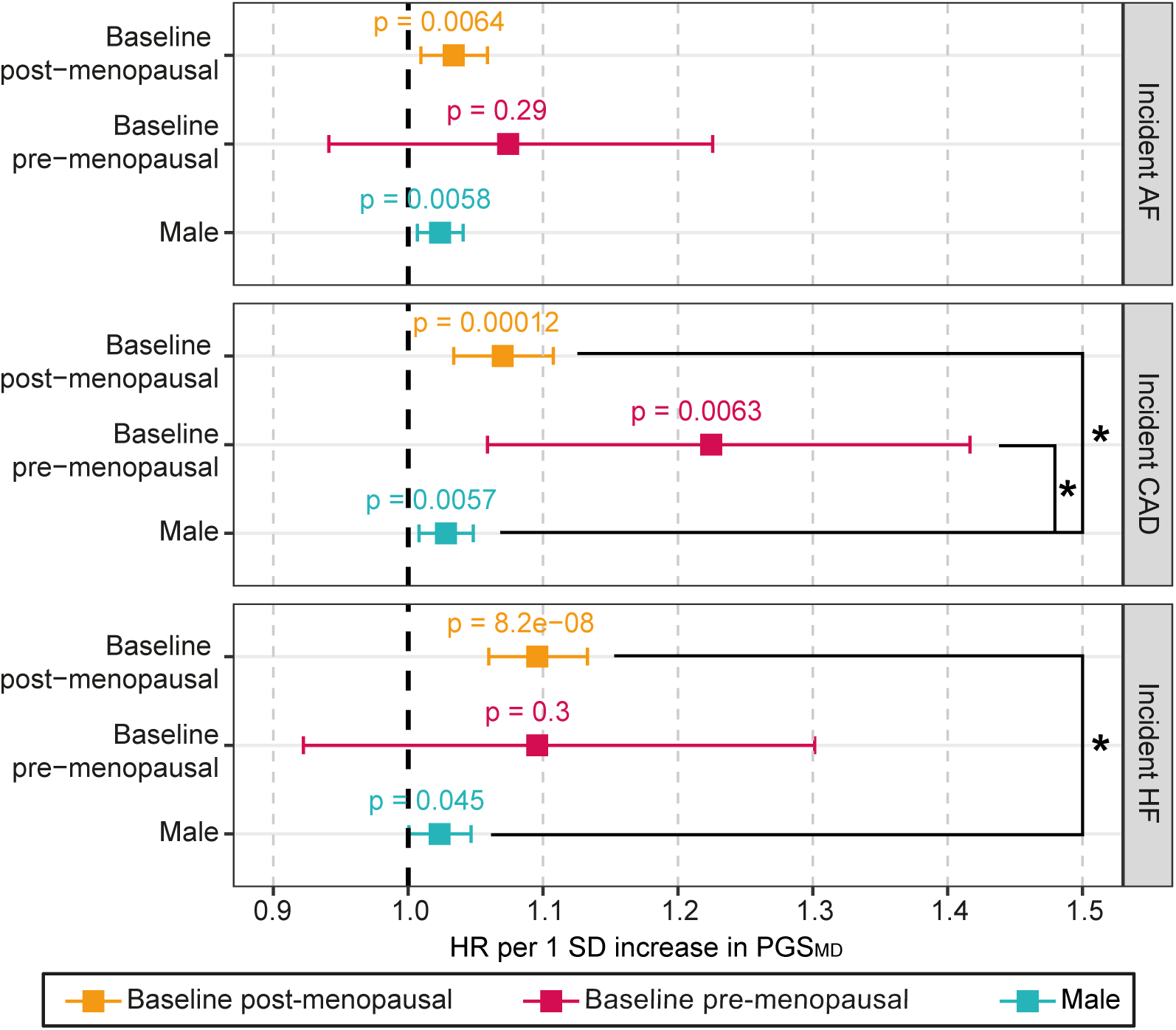
Age-specific change in risk of incident CVDs per SD increase in PGS_MD_ in the UK Biobank cohort, stratified by menopausal status. Associations for females who were baseline post-menopausal (orange) and baseline pre-menopausal (pink) were estimated with Cox proportional hazards regression models, and the estimates for the male cohort (blue) are shown for the purpose of comparison. X-axis shows the HR per SD increase in PGS, with p-values labelled, and error bars indicate 95% confidence intervals. Asterisk (*) indicates a statistically significant difference (two-sided Wald test p<0.05) in the beta values between the corresponding female cohort and male cohort. Dark grey line indicates HR of 1. AF, atrial fibrillation; CAD, coronary artery disease; HF, heart failure; HR, hazard ratio; MD, major depression; PGS, polygenic score; SD, standard deviation.

### The associations between the genetic risk of MD and CVDs are partly mediated by BMI, hypercholesterolemia, hypertension and smoking

To gain a mechanistic insight into the association between PGS_MD_ and CVD risk, we performed a sex-stratified mediation analysis to investigate whether traditional modifiable CVD risk factors could explain the observed sex difference. Although the association between PGS_MD_ and CVD risk in females was found to be partly mediated by a number of traditional risk factors (namely BMI, hypercholesterolemia, hypertension and smoking), given that the proportion of CVD risk explained by each of the risk factors was higher in males (though not all statistically significant after multiple testing correction), the observed higher risk in females is unlikely to be explained by these traditional risk factors (Table 2). For example, baseline BMI mediates 34% (95%CI: 21% - 78%; p=0.00060) of the association between PGS_MD_ and incident AF in males, but only 25% (95% CI: 18% - 41%; p<2E-16) in females (Table 2).

**Table 2.** Sex-stratified mediation analysis of risk factors modelled as mediators of the association between PGS_MD_ and incident CVDs in UK Biobank. Mediation effects that passed multiple testing correction (p<0.0017) are highlighted in bold.

| Outcome | Mediator | Sex | Prop. Mediated (average) | 95% CI Lower | 95% CI Upper | P |
| --- | --- | --- | --- | --- | --- | --- |
| Incident AF | Baseline BMI | Female | **0.25** | **0.18** | **0.41** | **< 2E-16** |
|  |  | Male | **0.34** | **0.21** | **0.78** | **0.0006** |
|  | Baseline hypercholesterolemia | Female | **0.085** | **0.038** | **0.25** | **0.0006** |
|  |  | Male | 0.13 | 0.043 | 0.43 | 0.0086 |
|  | Baseline hypertension | Female | **0.17** | **0.088** | **0.35** | **< 2E-16** |
|  |  | Male | 0.46 | 0.21 | 2.5 | 0.027 |
|  | Baseline smoking status | Female | **0.089** | **0.059** | **0.19** | **< 2E-16** |
|  |  | Male | 0.18 | 0.093 | 0.59 | 0.0062 |
|  | Baseline T2D | Female | 0.021 | 0.0072 | 0.08 | 0.012 |
|  |  | Male | 0.075 | 0.019 | 0.36 | 0.024 |
| Incident CAD | Baseline BMI | Female | **0.13** | **0.094** | **0.21** | **< 2E-16** |
|  |  | Male | 0.24 | 0.14 | 0.74 | 0.0064 |
|  | Baseline hypercholesterolemia | Female | **0.098** | **0.048** | **0.24** | **0.0004** |
|  |  | Male | 0.16 | 0.042 | 0.69 | 0.02 |
|  | Baseline hypertension | Female | **0.14** | **0.081** | **0.34** | **0.0002** |
|  |  | Male | 0.44 | 0.18 | 1.5 | 0.016 |
|  | Baseline smoking status | Female | **0.098** | **0.071** | **0.18** | **< 2E-16** |
|  |  | Male | 0.22 | 0.11 | 0.69 | 0.0074 |
|  | Baseline T2D | Female | 0.043 | 0.012 | 0.11 | 0.015 |
|  |  | Male | 0.072 | -0.018 | 0.39 | 0.09 |
| Incident HF | Baseline BMI | Female | **0.17** | **0.13** | **0.24** | **< 2E-16** |
|  |  | Male | 0.39 | 0.23 | 1.2 | 0.0034 |
|  | Baseline hypercholesterolemia | Female | **0.084** | **0.038** | **0.17** | **0.0006** |
|  |  | Male | 0.15 | 0.069 | 0.7 | 0.014 |
|  | Baseline hypertension | Female | **0.16** | **0.073** | **0.25** | **< 2E-16** |
|  |  | Male | 0.37 | 0.19 | 1.3 | 0.0078 |
|  | Baseline smoking status | Female | **0.099** | **0.063** | **0.14** | **< 2E-16** |
|  |  | Male | 0.31 | 0.15 | 1.1 | 0.016 |
|  | Baseline T2D | Female | 0.057 | 0.013 | 0.11 | 0.013 |
|  |  | Male | 0.22 | -0.77 | 1.6 | 0.079 |

### Genetic risk of MD is associated with increased CVD risk in females even in the absence of psychiatric diagnosis or psychiatric medication use

Amongst individuals who had no psychiatric disorder diagnosis and were not on any psychiatric medication (N=286,162), we observed a nominally positive association of PGS_MD_ with incident AF [HR_AF-female_ = 1.03 (95% CI: 1.01 - 1.06; p=0.013)], incident CAD [HR_CAD-female_ = 1.04 (95% CI: 1.01 - 1.08; p=0.016)] and incident HF [HR_HF-female_ = 1.07 (95% CI: 1.04 – 1.11; p=0.00012)] in females only (Figure 4).

**Figure 4.**
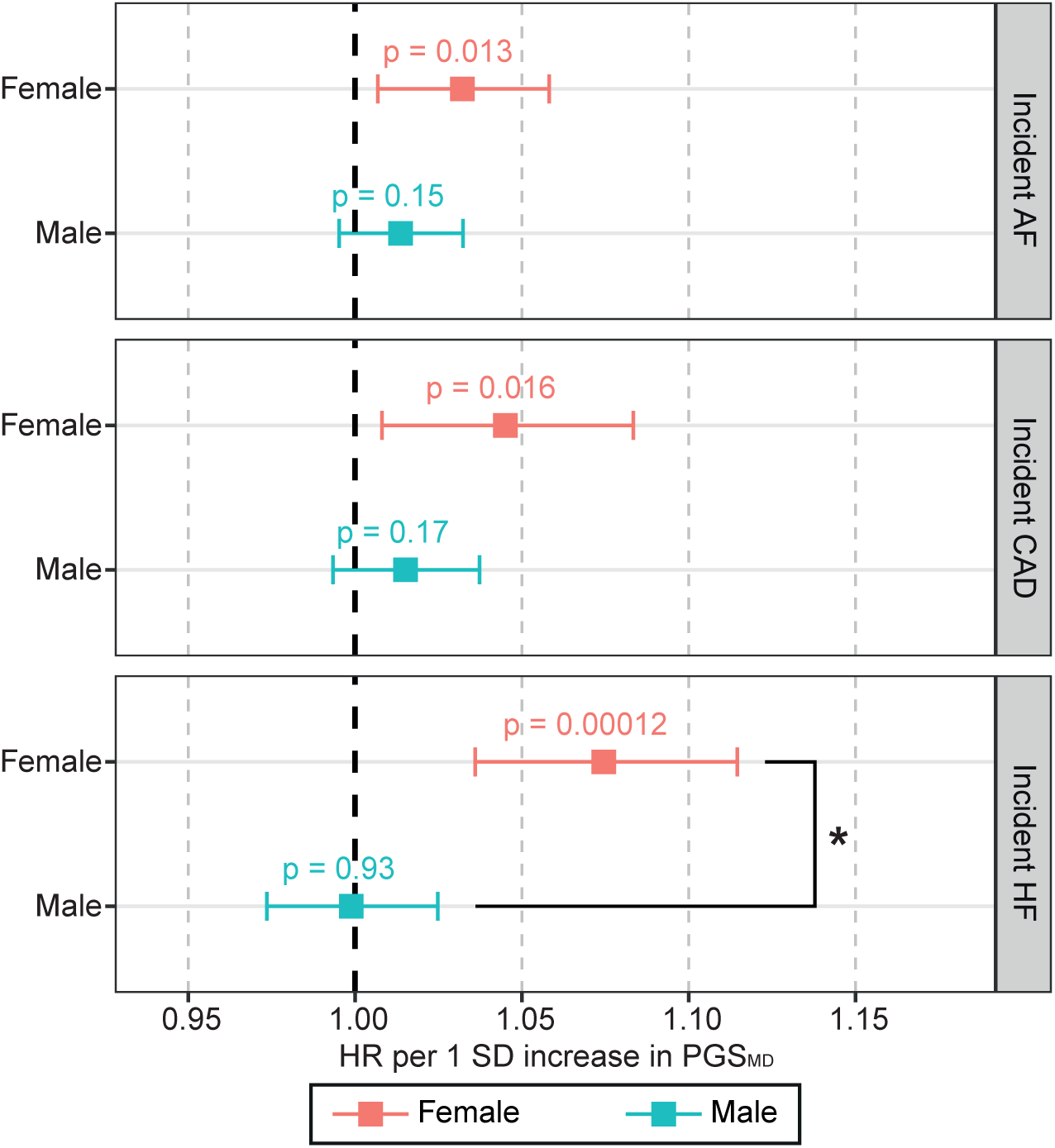
Age-specific change in risk of incident CVDs per SD increase in PGS_MD_ amongst UK Biobank individuals with no diagnosis of psychiatric disorders and psychiatric medications. Associations for the sex-stratified cohorts (red - female; blue - male) were estimated with Cox proportional hazards regression models. X-axis shows the HR per SD increase in PGS, with p-values labelled, and error bars indicate 95% confidence intervals. Asterisk (*) indicates a statistically significant difference in the beta values between females and males (two-sided Wald test p<0.05). Dark grey line indicates HR of 1. AF, atrial fibrillation; CAD, coronary artery disease; HF, heart failure; HR, hazard ratio; MD, major depression; PGS, polygenic score; SD, standard deviation.

### Replication of main findings in the BioVU cohort

We repeated our analysis in the independent BioVU cohort (N=49,057). Similar to the UK Biobank cohort, using a nominal significance threshold (p<0.05), we observed a positive association of PGS_MD_ with prevalent CAD in both sexes of the BioVU cohort [odds ratio (OR_CAD-female_) = 1.08 (95% CI: 1.03 – 1.12; p=0.00043) and OR_CAD-male_ = 1.04 (95% CI: 1.00 – 1.08; p=0.044)] (Supplementary Figure 2). However for HF, we observed a positive association in not only females [OR_HF-female_ = 1.07 (95% CI: 1.02 – 1.11; p=0.0036)], but also in males [OR_HF-male_ = 1.06 (95% CI: 1.02 – 1.10; p=0.0072)], with no significant difference in risk between the two sexes (two-sided Wald test p > 0.05) (Supplementary Figure 2). Unlike the UK Biobank, we did not observe an association between PGS_MD_ and AF in either sex. Interestingly, we observed a negative association of both PGS_BD_ and PGS_SCZ_ with prevalent CAD and HF in the males of BioVU, which may be suggestive of survivial bias (Supplementary Figure 2). A nominally significant genetic associations of PGS_MD_ with HF in both sexes, and with CAD in females was observed in the sensitivity analyses adjusting for the diagnosis of psychiatric disorders and antidepressant medication use (Supplementary Figure 3).

## Discussion

Using data from the UK Biobank cohort, we show for the first time the association between genetic predisposition to MD with incident AF and HF in females but not in males, and validate a previously reported sex difference (increased risk in women) in the association with CAD. As we observed a larger number of CVD cases in males and therefore had greater power to detect a risk estimate equivalent to that found in females, the observed sex difference in the depression-CVD associations was unlikely to be driven by differences in statistical power. We also show for the first time that the genetic risk of depression is associated with increased risks of CVDs even in the absence of psychiatric diagnosis or psychiatric medication use, and this association is thus not simply a consequence of behavioural changes or medication following a depression diagnosis. Interestingly, the association with incident CAD was consistent amongst females who were pre-menopausal at baseline (average age of 44.9 years at baseline) as well as in older females who were post-menopausal at baseline (mean age of 60.8 years at baseline), while the increased risk of incident AF and HF was only observed in the latter group. Our results from mediation analysis suggest that although the depression-CVD link is partly mediated by traditional CVD risk factors, these do not explain the sex difference observed in UK Biobank. Additional risk factors need to be investigated to understand the driver of the higher CVD risks in women.

We repeated these analyses in the independent BioVU cohort, which was limited by its much smaller sample size, potential ascertainment bias (data only available on individuals who were admitted to VUMC) and potential survival bias (incident data unavailable). We were able to replicate the association between the genetic risk of MD and increased HF and CAD risks in females, though both associations were also significant in males of BioVU, possibly due to the aforementioned limitations.

A few observational studies have found depression to be associated with increased AF, CAD and HF risk in women, and such associations were either weaker or absent in men ^12,13,32^. However, observational studies are prone to unmeasured confounder bias and reverse causation. These studies also have not been able to determine if this risk is independent of psychiatric medication use. Previous genetic analyses that utilise GWAS summary data to assess causality have shown that an increased genetic risk for MD is associated with an increased CAD and HF risk ^16^. However, these studies lacked a sex perspective, except for a previously reported genetic association between MD PGS and CAD observed in females but not males in a (smaller) BioVU cohort ^17^.

In this study, we observed comparable risk estimates between PGS_MD_ and incident CVDs amongst two cohorts of women who were at different menopausal stages at enrolment into the UK Biobank, although the risk estimates in the pre-menopausal cohort were restricted by sample size and thus did not reach statistical significance for AF and HF. The menopause transition involves extensive changes in sex hormones, body composition, and lipid profiles, which can increase CVD risk in women post-menopause ^15^. Our findings demonstrate that depression may be an important consideration in CVD risk assessment regardless of menopausal stage.

Mediation analyses suggest that the higher CVD risk associated with higher genetic predisposition to MD in females versus males is not explained by traditional CVD risk factors. Possible mechanisms underlying the sex difference in the comorbidity of depression and CVDs include hormonal dysregulation and pro-inflammatory responses. Specific subtypes of depression, such as postpartum depression and post-menopausal depression, suggest the involvement of hormone fluctuations in depression amongst females ^33^. Interestingly, Takotsubo cardiomyopathy, often triggered by emotional or physical stress, is more common in post-menopausal women (90% of cases) ^34^. At the same time, elevated inflammatory biomarkers, such as interleukin-6 and tumour necrosis factor alpha, are reportedly higher in depressed women relative to depressed men, suggesting a sex-differential inflammatory response to MD ^33^. These pro-inflammatory cytokines are linked to CVD risks ^35^. Further studies are required to understand the role of these factors in the heightened risk of CVDs amongst females with depression.

Currently, QRISK3 prediction algorithm (UK) is the only CVD risk prediction calculator that incorporates diagnoses of severe mental illnesses as risk factors for primary prevention ^36^. In New Zealand, individuals with severe mental illness are advised to undergo earlier and more frequent risk assessments ^37^. In Australia, severe mental illnesses are recommended as a ‘reclassification factor’ and to be used to refine CVD risk categorisation for individuals whose risk predicted from traditional risk factors lies close to a threshold of a higher-risk group ^38^. Furthermore, the American Heart Association (AHA) has recommended incorporation of a depression diagnosis as a risk factor of adverse prognosis amongst patients with acute coronary syndrome ^39^, but the diagnosis of depression is not currently incorporated into the calculation of CVD risk (Pooled Cohort Equations) in US ^40^. Given the observation of higher CVD risk in individuals who are genetically predisposed to depression, even in the absence of a depression diagnosis, future studies investigating any benefit of including such information in risk tools is warranted, especially in women.

The strengths of our study lie, firstly, in the investigation of sex-specific psychiatric-CVD comorbidity, and the ability to dissociate the effects of depression from medications and behaviour changes following diagnosis. However, limitations of our study need to be acknowledged. We derived PGSs from sex-combined summary statistics of GWAS, which might fail to capture genetic variants that confer sex-differential associations. The between-sex genetic correlations for BD, MD and SCZ are reported to range between 0.86 and 1, indicating moderate to subtle sex differences in the genetic architecture of these psychiatric disorders ^41^. Nevertheless, PGSs derived from sufficiently powered sex-stratified GWAS studies, which are becoming increasingly available for neuropsychiatric and cardiovascular traits ^41,42^, will provide additional information on the sex-differential genetic factors in diseases. Moreover, due to the lower number of CVD cases amongst females in UK Biobank, our analysis in the female cohort had lower power. In addition, the analysis of CVD risk difference between different menopausal states did not account for the potential effects of any menopause treatments which are known to impact CVD risks ^43^. Over 47.5% (54,350 out of 114,375) of baseline post-menopausal females in this study self-reported having used hormone replacement therapy at baseline (versus only 1.5% of the baseline pre-menopausal cohort). This is an important consideration for future studies to dissociate the effects of such treatments on the CVD risk amongst individuals with a genetic predisposition of MD. Furthermore, the “healthy-volunteer bias” in UK Biobank is well documented, where individuals with severe mental illnesses are underrepresented compared to the general population ^44^. While an underrepresentation of depression is also present in this cohort, it is estimated that this bias is likely more prominent for more several mental illnesses, namely schizophrenia ^44^.

To our best knowledge, this is the first study that has explored sex-specific association between the genetic risk of three different psychiatric disorders and risks of AF, CAD and HF, with further stratification for menopause status. Given that these CVDs have both shared and unique underlying disease biology, future research into the shared and unique biological pathways through which increased risk of depression increases the risk of different CVDs may be important for informing prevention strategies. CVD risk calculators that do not incorporate psychiatric disorders as a predictor are reported to underestimate CVD risk by 30% and 60% in men and women respectively ^45^. Our results warrant further investigation into the predictive value of depression risk in CVD risk calculators, even in the absence of a depression diagnosis, as well as further studies on how AF, CAD and HF risk differs across different menopausal stages.

## Data Availability

All data produced in the present work are contained in the manuscript. Publicly available GWAS summary statistics analysed in this study are properly referenced in the manuscript. The full GWAS summary statistics for the 23andMe discovery dataset will be made available through 23andMe to qualified researchers under an agreement with 23andMe that protects the privacy of the 23andMe participants. Please visit https://research.23andme.com/collaborate/#dataset-access/ for more information and to apply to access the data.

https://github.com/CNSGenomics/CVD_psych_PRS_Jiang

## Acknowledgments

We would like to thank the research participants and employees of 23andMe, inc. for making this work possible. The full GWAS summary statistics for the 23andMe discovery dataset will be made available through 23andMe to qualified researchers under an agreement with 23andMe that protects the privacy of the 23andMe participants. Please visit https://research.23andme.com/collaborate/#dataset-access/ for more information and to apply to access the data.

The dataset(s) used for the BioVU analyses described were obtained from Vanderbilt University Medical Center’s BioVU which is supported by numerous sources: institutional funding, private agencies, and federal grants. These include the NIH funded Shared Instrumentation Grant S10RR025141; and CTSA grants UL1TR002243, UL1TR000445, and UL1RR024975. Genomic data are also supported by investigator-led projects that include U01HG004798, R01NS032830, RC2GM092618, P50GM115305, U01HG006378, U19HL065962, R01HD074711; and additional funding sources listed at https://victr.vumc.org/biovu-funding/. This publication (journal article, etc.) was supported by Grant No. R01 HG011405, partially funded by the Office of Research on Women’s Health, Office of the Director, NIH and the National Human Genome Research Institute. Its contents are solely the responsibility of the authors and do not necessarily represent the official views of the Office of Research on Women’s Health or the National Human Genome Research Institute.

The scripts used in the study are available on: https://github.com/CNSGenomics/CVD_psych_PRS_Jiang.

## Non-Standard Abbreviations and Acronyms

AF: Atrial fibrillation
AHA: American Heart Association
BD: Bipolar disorder
BMI: Body mass index
CAD: Coronary artery disease
CI: Confidence interval
CPT: Current Procedural Terminology
CVD: Cardiovascular disease
EHR: Electronic health record
GWAS: Genome-wide association study
HF: Heart failure
HR: Hazard ratio
ICD: International Classification of Diseases
LD: Linkage disequilibrium
MD: Major depression
MR: Mendelian randomisation
OR: Odds ratio
PC: Principal component
PGS: Polygenic score
PRS-CS: Polygenic Risk Score–Continuous Shrinkage
SCZ: Schizophrenia
SD: Standard deviation
SNP: Single nucleotide polymorphism
T2D: Type II diabetes
VUMC: Vanderbilt University Medical Center

## Supplementary Material

## Supplementary Methods

### CVD and risk factor phenotype ascertainment in UK Biobank

AF cases were defined by the presence of self-reported atrial fibrillation or atrial flutter, self-reported cardioversion operation, ICD-10 (I48, I48.0-4, I48.9) or ICD-9 (4273) billing codes indicative of atrial fibrillation or atrial flutter, OPCS4 procedures (K57.1, K62.1-4), or death causes indicative of atrial fibrillation or atrial flutter (same as the ICD-10 codes).

CAD cases were ascertained based on myocardial infarction or coronary revascularisation. Myocardial infarction was defined by self-reported heart attack/myocardial infarction, or ICD-10 (I21.X, I22.X, I23.X, I24.1, I25.2) or ICD-9 (4109, 4119, 4129) billing codes for myocardial infarction. Coronary revascularisation was defined by OPCS4 codes for coronary artery bypass grafting (K40.1–4, K41.1–4, or K45.1–5) or coronary angioplasty with or without stenting (K49.1– 2, K49.8–9, K50.2, K75.1–4, K75.8–9).

HF cases were ascertained by the presence of self-reported heart failure/pulmonary odema or cardiomyopathy, ICD-10 (I11.0, I13.0, I13.2, I25.5, I42.0, I42.5, I42.8, I42.9, I50.0, I50.1, I50.9) or ICD-9 (4254, 4280, 4281, 4289) billing codes indicative of heart/ventricular failure or cardiomyopathy, or death causes indicative of heart/ventricular failure or cardiomyopathy. Individuals with hypertrophic cardiomyopathy, defined by self-reported illness, ICD-10 (I42.1, I42.2) billing codes, ICD-9 (4251) billing codes or death causes, were excluded from the HF analyses.

Hypercholesterolemia cases were defined by self-reported high cholesterol, ICD-10 (E780) or ICD-9 (27200, 27209) codes for hypercholesterolemia, or death causes indicative of hypercholesterolemia. Individuals who were not defined as a hypercholesterolemia case, but self-reported to be on cholesterol-lowering medication were removed from the hypercholesterolemia controls.

Hypertension cases were defined by self-reported hypertension, ICD-10 (I10) or ICD-9 (4010, 4011, 4019) codes for essential (primary) hypertension, or death causes indicative of hypertension. Individuals who were not defined as a hypertension case but self-reported to be taking blood pressure medications were removed from the hypertension controls.

T2D cases were defined by self-reported T2D, ICD-10 (E11.X) or ICD-9 (25000, 25010) codes for non-insulin-dependent diabetes mellitus, or death causes indicative of T2D. Individuals who were not defined as a hypertension case but had Type I diabetes or were taking insulin were removed from the T2D controls. Type I diabetes cases were defined by self-reported, ICD-10 (E10.X) codes, ICD-9 (25001, 25011) codes, or death causes.

### Psychiatric phenotype definitions in UK Biobank

BD cases were ascertained by the presence of ICD-10 (F30.0, F30.1, F30.2, F30.8, F30.9, F31.X) or ICD-9 (2960, 2961, 2969) codes for manic episode or bipolar affective disorder. SCZ cases were defined using ICD-10 (F20.X, F21.X, F22.X, F23.X, F24.X, F25.X, F28.X, F29.X) or ICD-9 (2953, 2959) codes for schizophrenia, schizotypal and delusional disorders. MD cases were defined by ICD-10 (F32.X, F33.X, F34.X, F38.X, F39.X) or ICD-9 (3119, 2962) codes for depressive episode, recurrent depressive disorder, persistent mood (affective) disorders, other mood (affective) disorders or unknown mood (affective) disorders. Individuals were removed from the MD cases if they were a case for BD or SCZ, had self-reported schizophrenia (1289), had an ICD-10 code of multiple personality disorder (F44.8), or were taking antipsychotics. The controls for BD, SCZ and MD were individuals who had no diagnosis of psychiatric disorders and were not taking any psychiatric medication (see definitions in Methods).

## Supplementary Figures

**Supplementary Figure 1.**
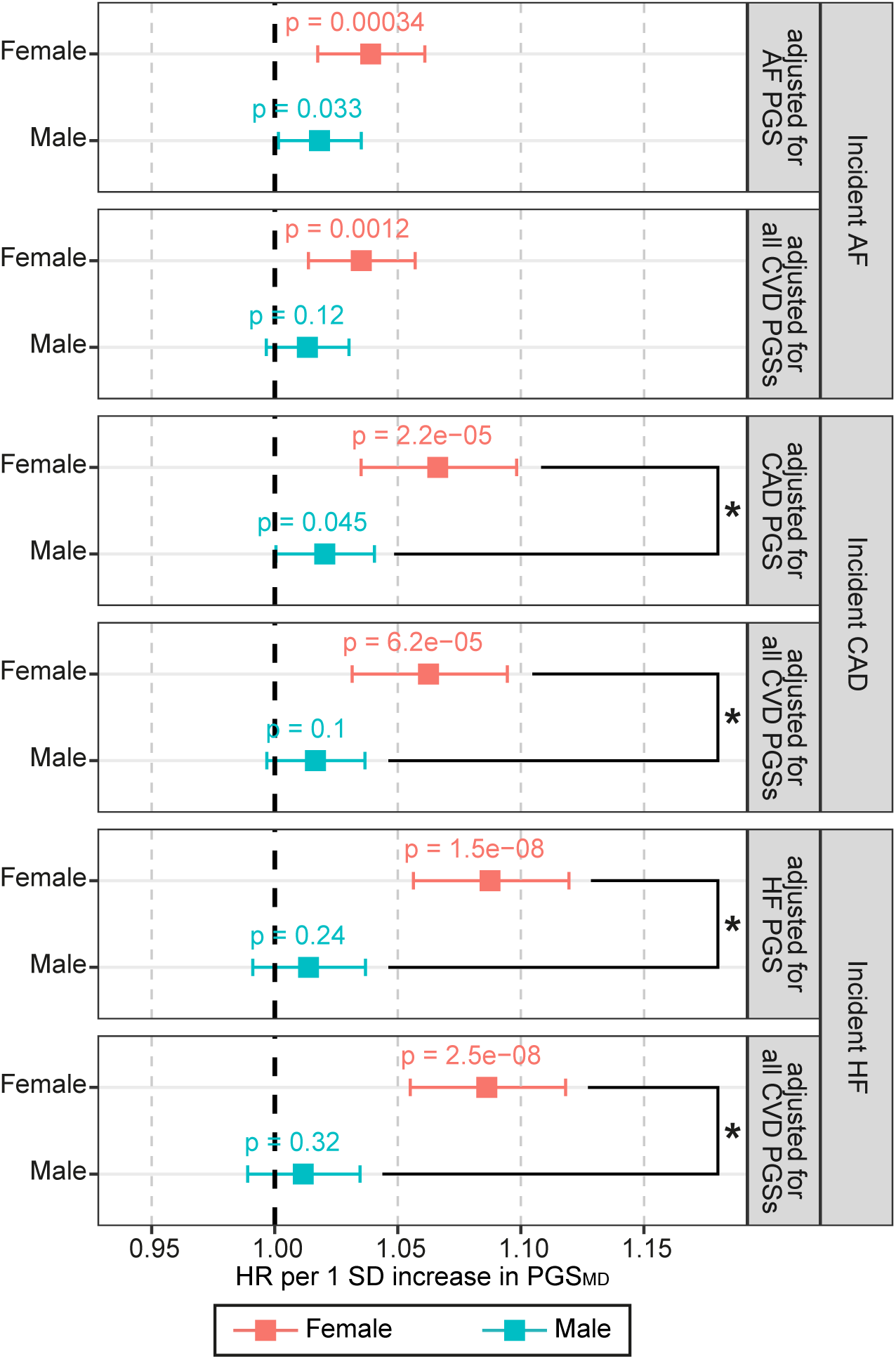
Sensitivity analyses on the association between PGS_MD_ and incident CVDs, performed on the UK Biobank cohort, where the corresponding CVD PGS or all three CVD PGSs (AF, CAD and HF) were added as covariates to the model. Associations for the sex-stratified cohorts (red - female; blue - male) were estimated with Cox proportional hazards regression models. X-axis shows the HR per SD increase in PGS, with p-values labelled, and error bars indicate 95% confidence intervals. Asterisk (*) indicates a statistically significant difference in the beta values between females and males (two-sided Wald test p<0.05). Dark grey line indicates HR of 1. AF, atrial fibrillation; CAD, coronary artery disease; CVD, cardiovascular disease; HF, heart failure; HR, hazard ratio; MD, major depression; PGS, polygenic score; SD, standard deviation.

**Supplementary Figure 2.**
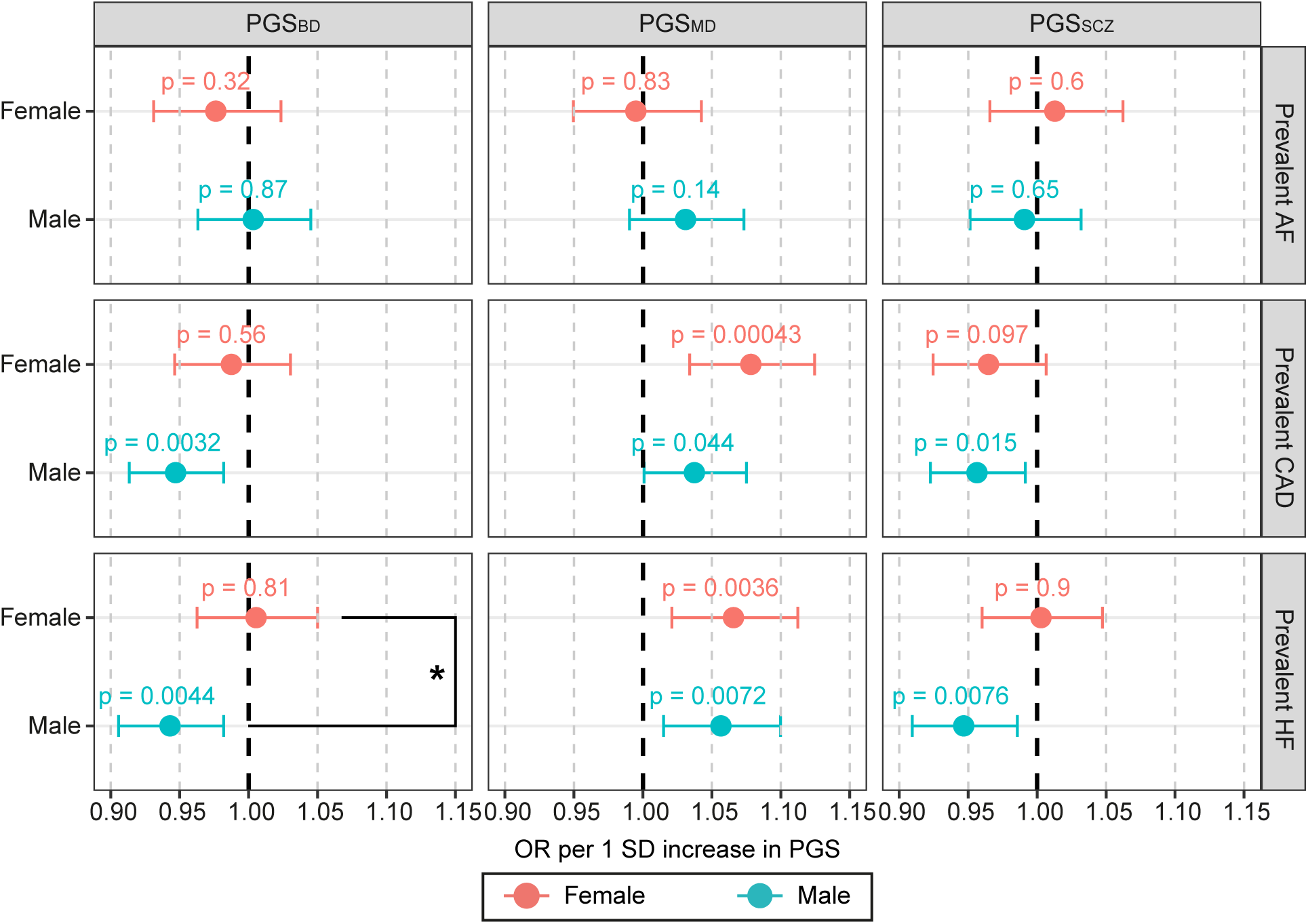
Association between psychiatric disorder PGSs and prevalent CVDs in the BioVU cohort. Associations for the sex-stratified cohorts (red - female; blue - male) were estimated with logistic regression. X-axis shows the OR per SD increase in PGS, with p-values labelled, and error bars indicate 95% confidence intervals. Asterisk (*) indicates a statistically significant difference in the beta values between females and males (two-sided Wald test p<0.05). Dark grey line indicates OR of 1. AF, atrial fibrillation; BD, bipolar disorder; CAD, coronary artery disease; HF, heart failure; MD, major depression; OR, odds ratio; PGS, polygenic score; SCZ, schizophrenia; SD, standard deviation.

**Supplementary Figure 3.**
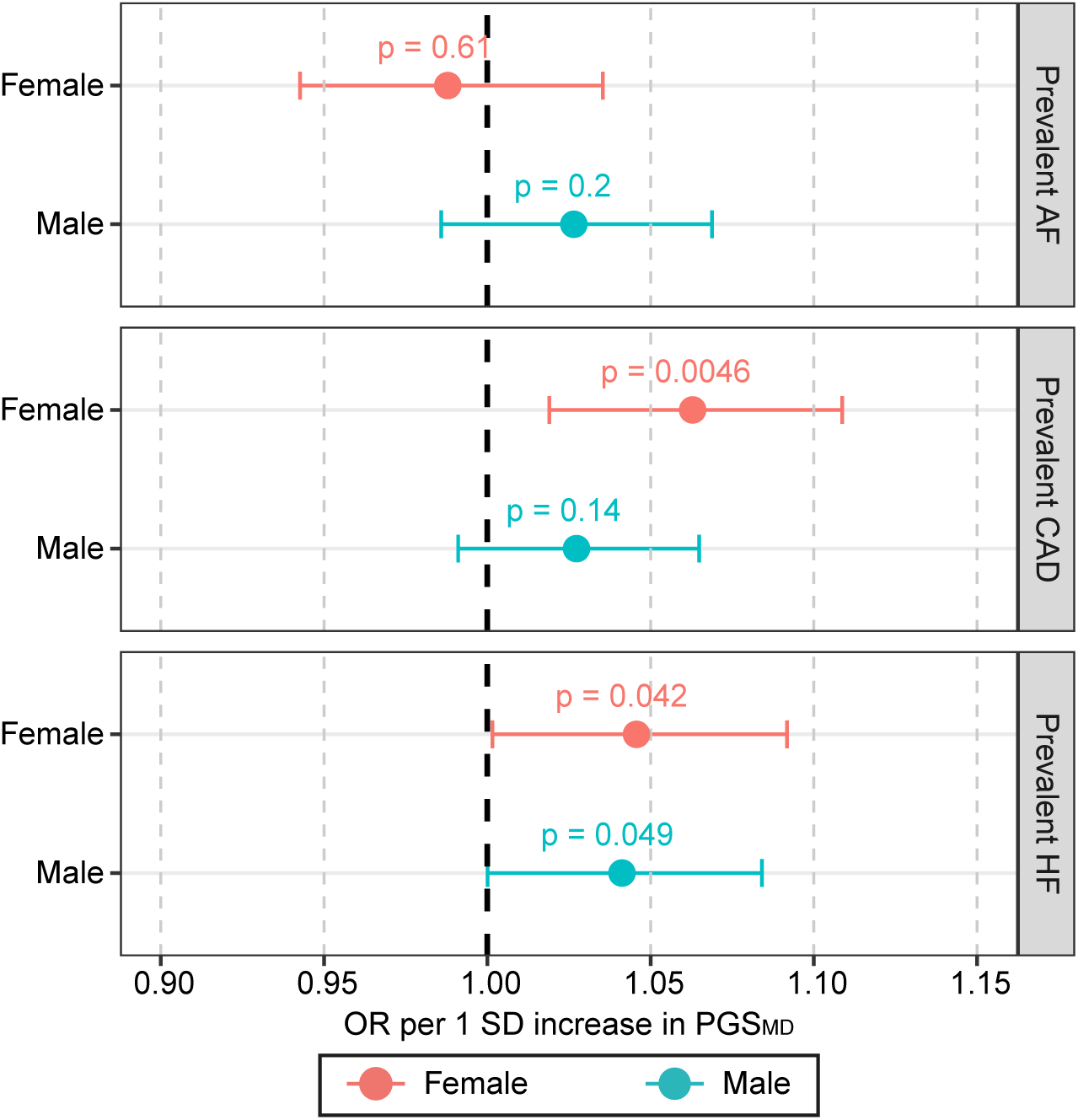
Association between PGS_MD_ and prevalent CVDs amongst individuals of the BioVU cohort, where the status of psychiatric disorders and antidepressant were added as covariates to the model. OR was estimated with logistic regression. X-axis shows the OR per SD increase in PGS, with p-values labelled, and error bars indicate 95% confidence intervals. Dark grey line indicates OR of 1. AF, atrial fibrillation; CAD, coronary artery disease; HF, heart failure; MD, major depression; OR, odds ratio; PGS, polygenic score; SD, standard deviation.

## Supplementary Tables

**Supplementary Table 1.**
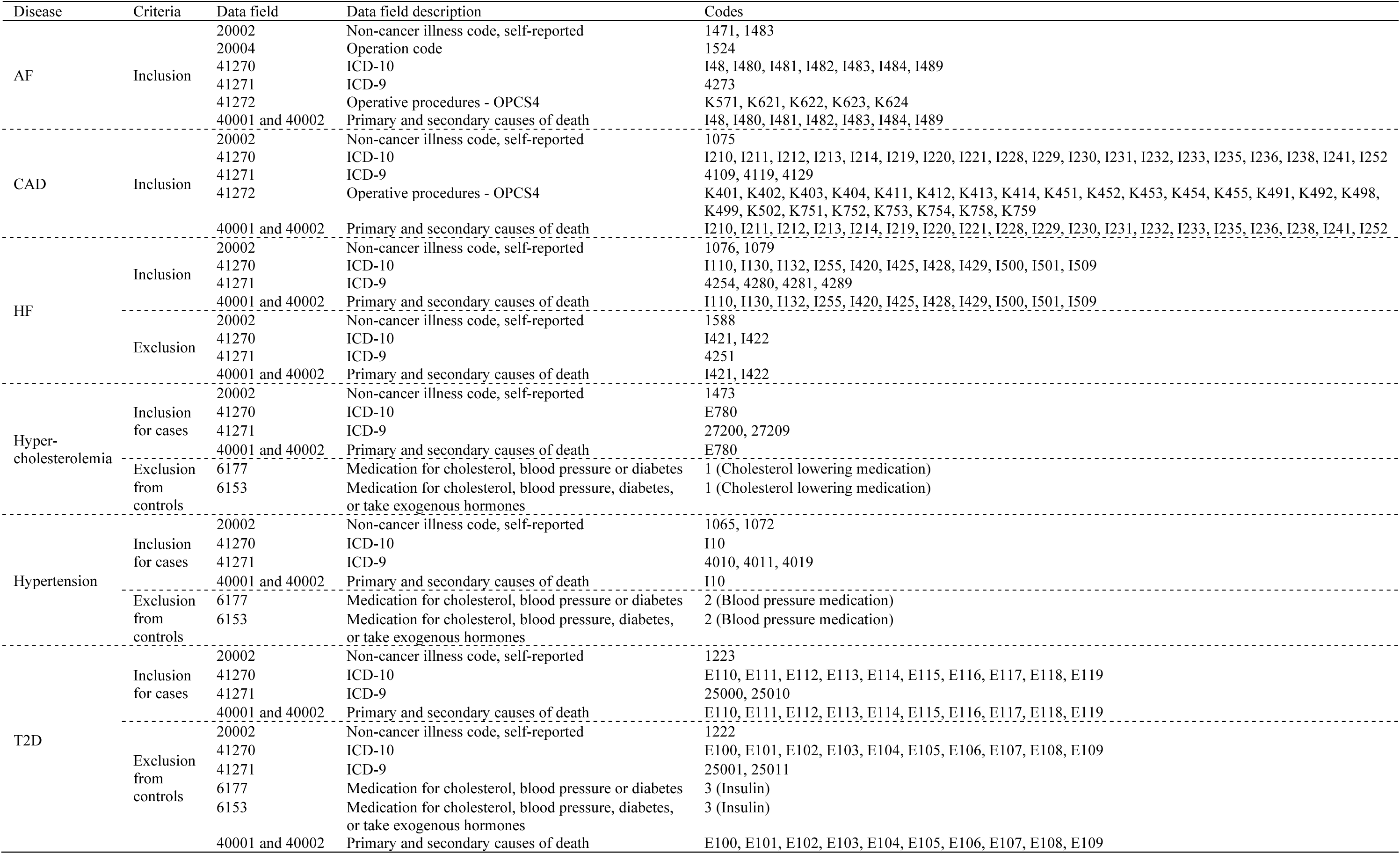
UK Biobank CVD and associated risk factors phenotype ascertainment criteria.

**Supplementary Table 2.**
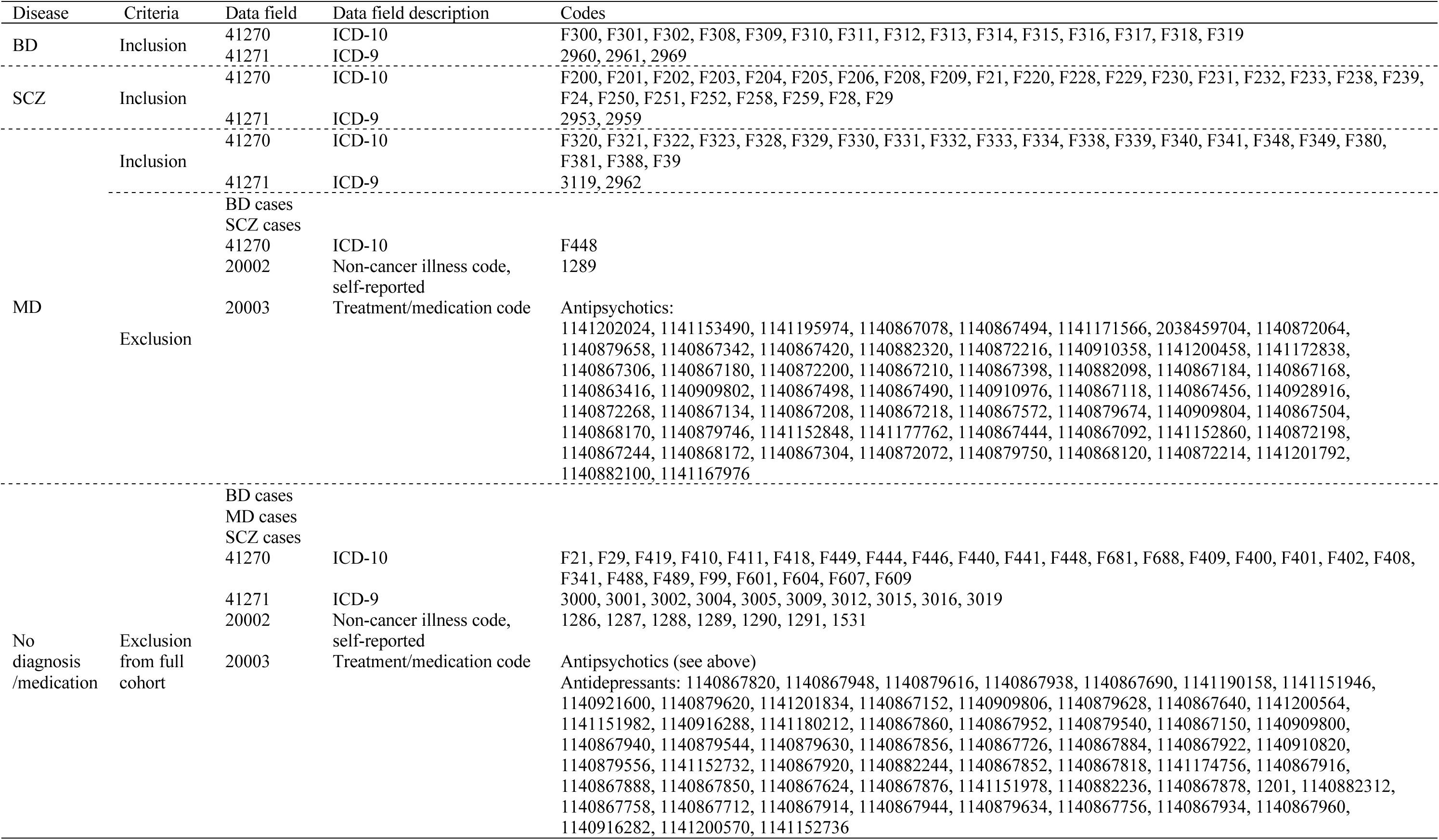
UK Biobank psychiatric disorder phenotype ascertainment criteria.

**Supplementary Table 3.** Summary of characteristics of participants in the UK Biobank cohort who had no diagnosis of psychiatric disorders and were not on any antidepressants or antipsychotics.

|  |  | Female |  | Male |  |
| --- | --- | --- | --- | --- | --- |
| Total Number |  | 148856 |  | 137306 |  |
| Average age at baseline in years (SD) |  | 57 (7.9) |  | 57.4 (8.1) |  |
| Average BMI at baseline (SD) |  | 26.7 (4.9) |  | 27.7 (4.1) |  |
| Current or previous smokers at baseline (%) |  | 59328 (39.9) |  | 68777 (50.1) |  |
|  |  | Case | Control | Case | Control |
| Incident AF | N (%) | 6375 (4.3) | 141290 (94.9) | 11643 (8.5) | 122363 (89.1) |
|  | Age at baseline (SD) | 62.7 (5.8) | 56.7 (7.9) | 62.3 (6.1) | 56.8 (8.1) |
| Incident CAD | N (%) | 3039 (2) | 144506 (97.1) | 8365 (6.1) | 122328 (89.1) |
|  | Age at baseline (SD) | 61.9 (6.2) | 56.9 (7.9) | 60.6 (6.9) | 56.9 (8.1) |
| Incident HF | N (%) | 2958 (2) | 145465 (97.7) | 5938 (4.3) | 130027 (94.7) |
|  | Age at baseline (SD) | 63.2 (5.7) | 56.9 (7.9) | 62.7 (6.1) | 57.1 (8.1) |
| Hypercholesterolemia status at baseline | N (%) | 14309 (9.6) | 121454 (81.6) | 22797 (16.6) | 93255 (67.9) |
|  | Age at baseline (SD) | 62.1 (5.7) | 56 (7.9) | 61.6 (6.3) | 55.7 (8.2) |
| Hypertension status at baseline | N (%) | 32739 (22) | 101591 (68.2) | 41542 (30.3) | 76756 (55.9) |
|  | Age at baseline (SD) | 60.6 (6.7) | 55.4 (7.9) | 60.5 (6.9) | 55.2 (8.2) |
| T2D status at baseline | N (%) | 1694 (1.1) | 141370 (95) | 3556 (2.6) | 123883 (90.2) |
|  | Age at baseline (SD) | 61.3 (6.6) | 56.8 (7.9) | 61.9 (6.1) | 57.1 (8.2) |

**Supplementary Table 4.** BioVU phenotype ascertainment criteria.

| Disease | Phecodes |
| --- | --- |
| AF | 427.2 |
| CAD | 411.4 |
| HF | 428, 428.1, 428.2, 428.3, 428.4 |
| BD | 296.1 |
| MD | 296.2, 296.22 |
| SCZ | 295.1 |
| Anxiety, dissociative and somatoform disorders | 300 |
| Personality disorders | 301 |
| Mood disorders | 296 |

**Supplementary Table 5.** Summary of characteristics in the BioVU cohort.

|  |  | Female |  | Male |  |
| --- | --- | --- | --- | --- | --- |
| Total Number |  | 28094 |  | 20963 |  |
| Median age in years (SD) |  | 26.76 (7.55) |  | 27.34 (6.60) |  |
| Median BMI (SD) |  | 51.00 (20.80) |  | 56.00 (22.50) |  |
| Tobacco Use Disorder (%) |  | 3129 (11.14) |  | 3486 (16.63) |  |
| Antidepressant use (%) |  | 16295 (58) |  | 9063 (43.23) |  |
| Psychiatric conditions (%) |  | 8993 (32.01) |  | 5043 (24.06) |  |
|  |  | Case | Control | Case | Control |
| AF | N (%) | 2256 (8.03) | 25838 (91.97) | 3416 (16.30) | 17547 (83.70) |
|  | Median age (SD) | 72.00 (12.62) | 49.00 (20.392) | 69.00 (12.33) | 53.00 (22.6) |
| CAD | N (%) | 2881 (10.25) | 25213 (97.1) | 5487 (26.17) | 15476 (73.83) |
|  | Median age (SD) | 68 (12.23) | 49.00 (20.58) | 68.00 (10.95) | 50.00 (22.84) |
| HF | N (%) | 2514 (8.95) | 25580 (91.05) | 3183 (15.18) | 17,780 (84.82) |
|  | Median age (SD) | 67.00 (18.03) | 50.00 (20.53) | 67.00 (16.65) | 54.0 (22.61) |

**Supplementary Table 6.**
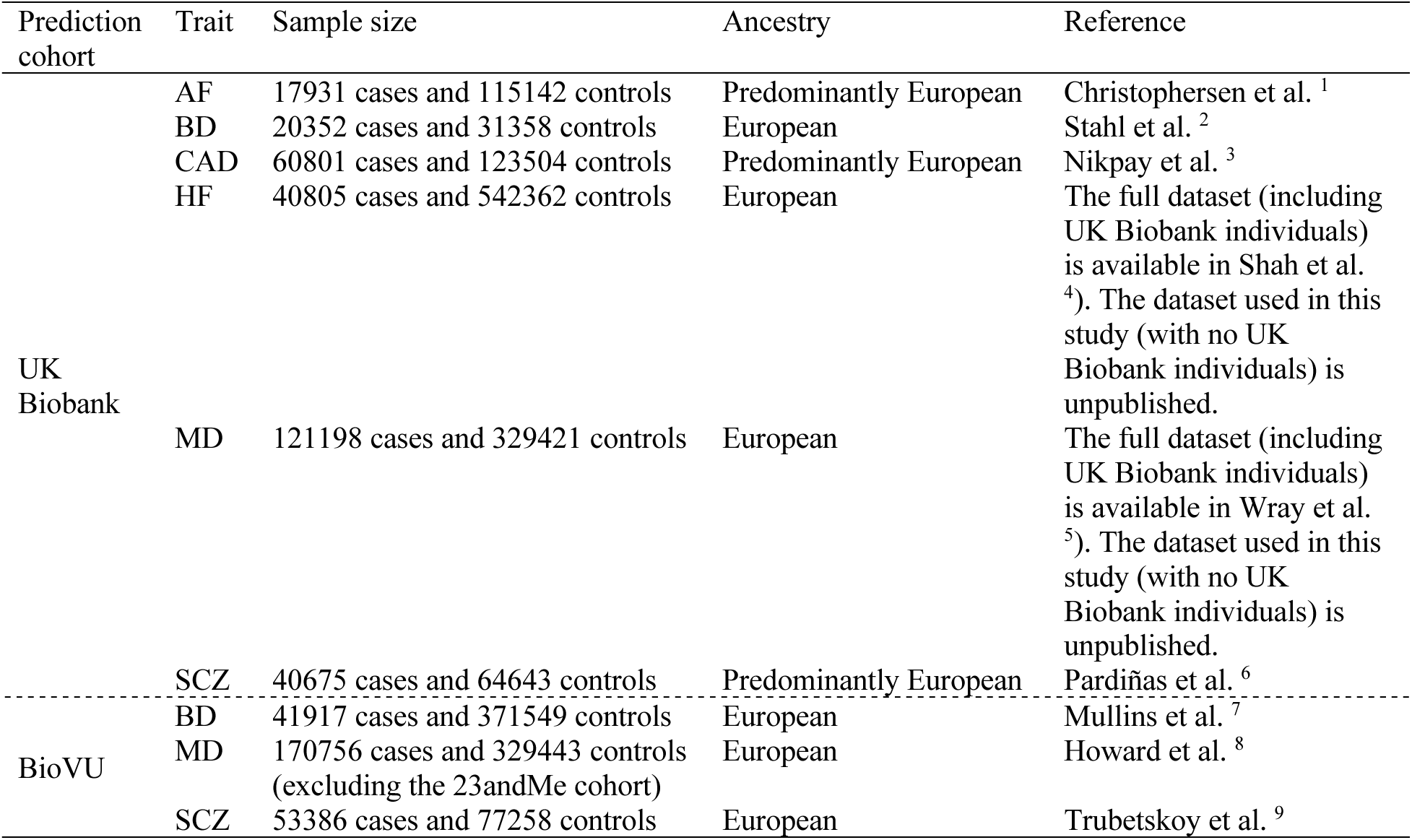
Sources of GWAS summary statistics used in PGS SNP weight estimation for UK Biobank and BioVU cohorts.

**Supplementary Table 7.**
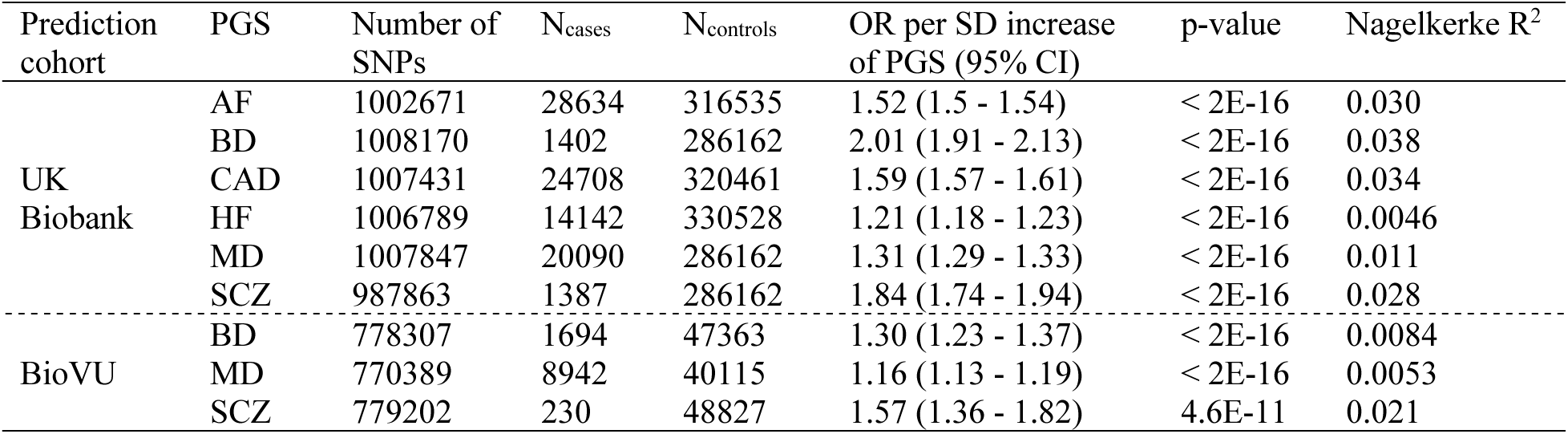
Prediction accuracy of PGSs generated for the UK Biobank cohort.

| Prediction cohort | PGS | Number of SNPs | N<sub>cases</sub> | N<sub>controls</sub> | OR per SD increase of PGS (95% CI) | p-value | Nagelkerke R<sup>2</sup> |
| --- | --- | --- | --- | --- | --- | --- | --- |
| UK Biobank | AF | 1002671 | 28634 | 316535 | 1.52 (1.5 - 1.54) | < 2E-16 | 0.030 |
|  | BD | 1008170 | 1402 | 286162 | 2.01 (1.91 - 2.13) | < 2E-16 | 0.038 |
|  | CAD | 1007431 | 24708 | 320461 | 1.59 (1.57 - 1.61) | < 2E-16 | 0.034 |
|  | HF | 1006789 | 14142 | 330528 | 1.21 (1.18 - 1.23) | < 2E-16 | 0.0046 |
|  | MD | 1007847 | 20090 | 286162 | 1.31 (1.29 - 1.33) | < 2E-16 | 0.011 |
|  | SCZ | 987863 | 1387 | 286162 | 1.84 (1.74 - 1.94) | < 2E-16 | 0.028 |
| BioVU | BD | 778307 | 1694 | 47363 | 1.30 (1.23 - 1.37) | < 2E-16 | 0.0084 |
|  | MD | 770389 | 8942 | 40115 | 1.16 (1.13 - 1.19) | < 2E-16 | 0.0053 |
|  | SCZ | 779202 | 230 | 48827 | 1.57 (1.36 - 1.82) | 4.6E-11 | 0.021 |

**Supplementary Table 8.** CVD incidence and average recruitment age of females who were baseline pre- and post-menopause in the UK Biobank (*** indicates a chi square p<2E-16 for comparison of CVD incidence between the two menopausal groups)

|  |  | Baseline pre-menopause |  | Baseline post-menopause |  |  |
| --- | --- | --- | --- | --- | --- | --- |
|  |  | Case | Control | Case | Control |  |
| AF | Count (%) | 222 (0.8) | 27076 (99.2) | 6849 (6.1) | 106288 (93.9) | *** |
|  | Average age at baseline in years (SD) | 45.3 (2.5) | 44.9 (2.6) | 63.6 (4.6) | 60.6 (5.4) |  |
| CAD | Count (%) | 185 (0.7) | 27103 (99.3) | 3288 (2.9) | 109621 (97.1) | *** |
|  | Average age at baseline in years (SD) | 45.4 (2.4) | 44.9 (2.6) | 62.9 (4.9) | 60.7 (5.4) |  |
| HF | Count (%) | 130 (0.5) | 27181 (99.5) | 3510 (3.1) | 110383 (96.9) | *** |
|  | Average age at baseline in years (SD) | 45.3 (2.4) | 44.9 (2.6) | 63.7 (4.8) | 60.7 (5.4) |  |

## References

1. Graboys TB. Stress and the aching heart. N Engl J Med. 1984;311:594–595.

2. Correll CU, Solmi M, Veronese N, Bortolato B, Rosson S, Santonastaso P, Thapa-Chhetri N, Fornaro M, Gallicchio D, Collantoni E, et al. Prevalence, incidence and mortality from cardiovascular disease in patients with pooled and specific severe mental illness: a large-scale meta-analysis of 3,211,768 patients and 113,383,368 controls. World Psychiatry. 2017;16:163–180.

3. Marano G, Traversi G, Romagnoli E, Catalano V, Lotrionte M, Abbate A, Biondi-Zoccai G, Mazza M. Cardiologic side effects of psychotropic drugs. J Geriatr Cardiol. 2011;8:243–253.

4. Larsen BA, Christenfeld NJS. Cardiovascular disease and psychiatric comorbidity: the potential role of perseverative cognition. Cardiovasc Psychiatry Neurol. 2009;2009:791017–791017.

5. Hasin DS, Sarvet AL, Meyers JL, Saha TD, Ruan WJ, Stohl M, Grant BF. Epidemiology of adult DSM-5 major depressive disorder and its specifiers in the United States. JAMA Psychiatry. 2018;75:336–346.

6. Marcus SM, Kerber KB, Rush AJ, Wisniewski SR, Nierenberg A, Balasubramani GK, Ritz L, Kornstein S, Young EA, Trivedi MH. Sex differences in depression symptoms in treatment-seeking adults: confirmatory analyses from the Sequenced Treatment Alternatives to Relieve Depression study. Compr Psychiatry. 2008;49:238–246.

7. Ochoa S, Usall J, Cobo J, Labad X, Kulkarni J. Gender differences in schizophrenia and first-episode psychosis: a comprehensive literature review. Schizophr Res Treatment. 2012;2012:916198.

8. Vega P, Barbeito S, de Azúa SR, Martínez-Cengotitabengoa M, González–Ortega I, Saenz M, González-Pinto A. Bipolar disorder differences between genders: special considerations for women. Womens Health. 2011;7:663–676.

9. Wilkins JT, Ning H, Berry J, Zhao L, Dyer AR, Lloyd-Jones DM. Lifetime risk and years lived free of total cardiovascular disease. JAMA. 2012;308:1795–1801.

10. Vogel B, Acevedo M, Appelman Y, Bairey Merz CN, Chieffo A, Figtree GA, Guerrero M, Kunadian V, Lam CSP, Maas AHEM, et al. The Lancet women and cardiovascular disease Commission: reducing the global burden by 2030. The Lancet. 2021;397:2385–2438.

11. Möller-Leimkühler AM. Gender differences in cardiovascular disease and comorbid depression. Dialogues Clin Neurosci. 2007;9:71–83.

12. Williams SA, Kasl SV, Heiat A, Abramson JL, Krumholz HM, Vaccarino V. Depression and risk of heart failure among the elderly: a prospective community-based study. Psychosom Med. 2002;64:6–12.

13. Shah AJ, Ghasemzadeh N, Zaragoza-Macias E, Patel R, Eapen DJ, Neeland IJ, Pimple PM, Zafari AM, Quyyumi AA, Vaccarino V. Sex and age differences in the association of depression with obstructive coronary artery disease and adverse cardiovascular events. J Am Heart Assoc. 2014;3:e000741.

14. Meng R, Yu C, Liu N, He M, Lv J, Guo Y, Bian Z, Yang L, Chen Y, Zhang X, et al. Association of depression with all-cause and cardiovascular disease mortality among adults in China. JAMA Network Open. 2020;3:e1921043–e1921043.

15. Khoudary SRE, Aggarwal B, Beckie TM, Hodis HN, Johnson AE, Langer RD, Limacher MC, Manson JE, Stefanick ML, Allison MA. Menopause transition and cardiovascular disease risk: implications for timing of early prevention: a scientific statement from the American Heart Association. Circulation. 2020;142:e506–e532.

16. Lu Y, Wang Z, Georgakis MK, Lin H, Zheng L. Genetic liability to depression and risk of coronary artery disease, myocardial infarction, and other cardiovascular outcomes. J Am Heart Assoc. 2021;10:e017986.

17. Dennis J, Sealock J, Levinson RT, Farber-Eger E, Franco J, Fong S, Straub P, Hucks D, Song WL, Linton MF, et al. Genetic risk for major depressive disorder and loneliness in sex-specific associations with coronary artery disease. Mol Psychiatry. 2021;26:4254–4264.

18. Bycroft C, Freeman C, Petkova D, Band G, Elliott LT, Sharp K, Motyer A, Vukcevic D, Delaneau O, O’Connell J, et al. The UK Biobank resource with deep phenotyping and genomic data. Nature. 2018;562:203–209.

19. Yengo L, Sidorenko J, Kemper KE, Zheng Z, Wood AR, Weedon MN, Frayling TM, Hirschhorn J, Yang J, Visscher PM, et al. Meta-analysis of genome-wide association studies for height and body mass index in ∼700000 individuals of European ancestry. Hum Mol Genet. 2018;27:3641–3649.

20. Carroll RJ, Bastarache L, Denny JC. R PheWAS: data analysis and plotting tools for phenome-wide association studies in the R environment. Bioinformatics. 2014;30:2375–2376.

21. Lloyd-Jones LR, Zeng J, Sidorenko J, Yengo L, Moser G, Kemper KE, Wang H, Zheng Z, Magi R, Esko T, et al. Improved polygenic prediction by Bayesian multiple regression on summary statistics. Nat Commun. 2019;10:5086.

22. Ni G, Zeng J, Revez JA, Wang Y, Zheng Z, Ge T, Restuadi R, Kiewa J, Nyholt DR, Coleman JRI, et al. A comparison of ten polygenic score methods for psychiatric disorders applied across multiple cohorts. Biol Psychiatry. 2021;90:611–620.

23. Chang CC, Chow CC, Tellier LCAM, Vattikuti S, Purcell SM, Lee JJ. Second-generation PLINK: rising to the challenge of larger and richer datasets. GigaScience. 2015;4:s13742-13015-10047-13748.

24. Ge T, Chen C-Y, Ni Y, Feng Y-CA, Smoller JW. Polygenic prediction via Bayesian regression and continuous shrinkage priors. Nat Commun. 2019;10:1776.

25. Mangiafico S. rcompanion: Functions to support extension education program evaluation. In; 2022.

26. Khera AV, Chaffin M, Aragam KG, Haas ME, Roselli C, Choi SH, Natarajan P, Lander ES, Lubitz SA, Ellinor PT, et al. Genome-wide polygenic scores for common diseases identify individuals with risk equivalent to monogenic mutations. Nat Genet. 2018;50:1219–1224.

27. Wang Y, Namba S, Lopera E, Kerminen S, Tsuo K, Läll K, Kanai M, Zhou W, Wu K-H, Favé M-J, et al. Global Biobank analyses provide lessons for developing polygenic risk scores across diverse cohorts. Cell Genomics. 2023;3:100241.

28. Reginsson GW, Ingason A, Euesden J, Bjornsdottir G, Olafsson S, Sigurdsson E, Oskarsson H, Tyrfingsson T, Runarsdottir V, Hansdottir I, et al. Polygenic risk scores for schizophrenia and bipolar disorder associate with addiction. Addict Biol. 2018;23:485–492.

29. Therneau TM. A Package for Survival Analysis in R. R package version 3.5–7, https://CRAN.R-project.org/package=survival. In; 2023.

30. Zhang F, Cao H, Baranova A. Shared genetic liability and causal associations between major depressive disorder and cardiovascular diseases. Front Cardiovasc Med. 2021;8.

31. Tingley D, Yamamoto T, Hirose K, Keele L, Imai K. mediation: R package for causal mediation analysis. Journal of Statistical Software. 2014;59:1–38.

32. Kim YG, Lee K-N, Han K-D, Han K-M, Min K, Choi HY, Choi YY, Shim J, Choi J-I, Kim Y-H. Association of depression with atrial fibrillation in South Korean adults. JAMA Network Open. 2022;5:e2141772–e2141772.

33. Tobaldini E, Carandina A, Toschi-Dias E, Erba L, Furlan L, Sgoifo A, Montano N. Depression and cardiovascular autonomic control: a matter of vagus and sex paradox. Neurosci Biobehav Rev. 2020;116:154–161.

34. Schneider B, Sechtem U. Influence of age and gender in Takotsubo Syndrome. Heart Fail Clin. 2016;12:521–530.

35. Kaptoge S, Seshasai SRK, Gao P, Freitag DF, Butterworth AS, Borglykke A, Di Angelantonio E, Gudnason V, Rumley A, Lowe GDO, et al. Inflammatory cytokines and risk of coronary heart disease: new prospective study and updated meta-analysis. Eur Heart J. 2014;35:578–589.

36. Hippisley-Cox J, Coupland C, Brindle P. Development and validation of QRISK3 risk prediction algorithms to estimate future risk of cardiovascular disease: prospective cohort study. BMJ. 2017;357:j2099.

37. Ministry of Health New Zealand. Cardiovascular disease risk assessment and management for primary care. 2018.

38. Commonwealth of Australia as represented by the Department of Health and Aged Care. Australian Guideline for assessing and managing cardiovascular disease risk. 2023.

39. Lichtman JH, Froelicher ES, Blumenthal JA, Carney RM, Doering LV, Frasure-Smith N, Freedland KE, Jaffe AS, Leifheit-Limson EC, Sheps DS, et al. Depression as a risk factor for poor prognosis among patients with acute coronary syndrome: systematic review and recommendations. Circulation. 2014;129:1350–1369.

40. Grundy SM, Stone NJ, Bailey AL, Beam C, Birtcher KK, Blumenthal RS, Braun LT, de Ferranti S, Faiella-Tommasino J, Forman DE, et al. 2018 AHA/ACC/AACVPR/AAPA/ABC/ACPM/ADA/AGS/APhA/ASPC/NLA/PCNA guideline on the management of blood cholesterol: a report of the American College of Cardiology/American Heart Association Task Force on Clinical Practice Guidelines. Circulation. 2019;139:e1082–e1143.

41. Martin J, Khramtsova EA, Goleva SB, Blokland GAM, Traglia M, Walters RK, Hübel C, Coleman JRI, Breen G, Børglum AD, et al. Examining sex-differentiated genetic effects across neuropsychiatric and behavioral traits. Biol Psychiatry. 2021;89:1127–1137.

42. Deloukas P, Kanoni S, Willenborg C, Farrall M, Assimes TL, Thompson JR, Ingelsson E, Saleheen D, Erdmann J, Goldstein BA, et al. Large-scale association analysis identifies new risk loci for coronary artery disease. Nat Genet. 2013;45:25–33.

43. Boardman HMP, Hartley L, Eisinga A, Main C, Roqué i Figuls M, Bonfill Cosp X, Gabriel Sanchez R, Knight B. Hormone therapy for preventing cardiovascular disease in post-menopausal women. Cochrane Database of Systematic Reviews. 2015.

44. Schoeler T, Speed D, Porcu E, Pirastu N, Pingault J-B, Kutalik Z. Participation bias in the UK Biobank distorts genetic associations and downstream analyses. Nature Human Behaviour. 2023;7:1216–1227.

45. Cunningham R, Poppe K, Peterson D, Every-Palmer S, Soosay I, Jackson R. Prediction of cardiovascular disease risk among people with severe mental illness: A cohort study. PLoS One. 2019;14:e0221521–e0221521.

## References

1. Christophersen IE, Rienstra M, Roselli C, Yin X, Geelhoed B, Barnard J, Lin H, Arking DE, Smith AV, Albert CM, et al. Large-scale analyses of common and rare variants identify 12 new loci associated with atrial fibrillation. Nat Genet. 2017;49:946–952.

2. Stahl EA, Breen G, Forstner AJ, McQuillin A, Ripke S, Trubetskoy V, Mattheisen M, Wang Y, Coleman JRI, Gaspar HA, et al. Genome-wide association study identifies 30 loci associated with bipolar disorder. Nat Genet. 2019;51:793–803.

3. Nikpay M, Goel A, Won H-H, Hall LM, Willenborg C, Kanoni S, Saleheen D, Kyriakou T, Nelson CP, Hopewell JC, et al. A comprehensive 1000 Genomes–based genome-wide association meta-analysis of coronary artery disease. Nat Genet. 2015;47:1121–1130.

4. Shah S, Henry A, Roselli C, Lin H, Sveinbjörnsson G, Fatemifar G, Hedman ÅK, Wilk JB, Morley MP, Chaffin MD, et al. Genome-wide association and Mendelian randomisation analysis provide insights into the pathogenesis of heart failure. Nat Commun. 2020;11:163.

5. Wray NR, Ripke S, Mattheisen M, Trzaskowski M, Byrne EM, Abdellaoui A, Adams MJ, Agerbo E, Air TM, Andlauer TMF, et al. Genome-wide association analyses identify 44 risk variants and refine the genetic architecture of major depression. Nat Genet. 2018;50:668–681.

6. Pardiñas AF, Holmans P, Pocklington AJ, Escott-Price V, Ripke S, Carrera N, Legge SE, Bishop S, Cameron D, Hamshere ML, et al. Common schizophrenia alleles are enriched in mutation-intolerant genes and in regions under strong background selection. Nat Genet. 2018;50:381–389.

7. Mullins N, Forstner AJ, O’Connell KS, Coombes B, Coleman JRI, Qiao Z, Als TD, Bigdeli TB, Børte S, Bryois J, et al. Genome-wide association study of more than 40,000 bipolar disorder cases provides new insights into the underlying biology. Nat Genet. 2021;53:817–829.

8. Howard DM, Adams MJ, Clarke TK, Hafferty JD, Gibson J, Shirali M, Coleman JRI, Hagenaars SP, Ward J, Wigmore EM, et al. Genome-wide meta-analysis of depression identifies 102 independent variants and highlights the importance of the prefrontal brain regions. Nat Neurosci. 2019;22:343–352.

9. Trubetskoy V, Pardiñas AF, Qi T, Panagiotaropoulou G, Awasthi S, Bigdeli TB, Bryois J, Chen C-Y, Dennison CA, Hall LS, et al. Mapping genomic loci implicates genes and synaptic biology in schizophrenia. Nature. 2022;604:502–508.

